# Explore-exploit instability reveals computational decision-making heterogeneity in early psychosis

**DOI:** 10.1101/2025.04.29.25326698

**Authors:** Cathy S. Chen, Evan Knep, Veldon-James Laurie, Olivia Calvin, R. Becket Ebitz, Melissa Fisher, Michael-Paul Schallmo, Scott R. Sponheim, Matthew V. Chafee, Sarah R. Heilbronner, Nicola M. Grissom, A. David Redish, Angus W. MacDonald, Sophia Vinogradov, Caroline Demro

## Abstract

**Background and Hypothesis:** Psychosis spectrum illnesses are characterized by impaired goal-directed behavior and significant clinical and neurophysiological heterogeneity. This study investigated cognitive heterogeneity by applying computational modeling to trial-wise decision making task behavior.

**Study Design:** 75 participants with Early Psychosis (EP) and 68 controls completed a dynamic decision-making task during two baseline sessions as part of a larger longitudinal fMRI study.

**Study Results:** Consistent with prior studies, EP exhibited more choice switching. However, this was not explained by reward learning deficits as no group difference in reward acquisition was found. Instead, a Hidden Markov model fit to choice sequences revealed increased exploration as a result of higher probability of transition from exploitation to exploration in EP, leaving a favorable option too soon. A Bayesian learner model that estimates both value and uncertainty characterized EP behavior better than traditional RL models with fixed learning rates. Results of computational modeling implicated elevated uncertainty sensitivity and decision noise as independent contributors to suboptimal transition into exploration among EP. Task strategy yielded three computational subtypes (normative, uncertainty-sensitive, high decision-noise) with unique cognitive and symptom profiles.

**Conclusions:** These specific microcognitive disruptions underlying the distinct neurocomputational subtypes are individually measurable and may have the potential for targeted interventions.

## Introduction

Psychosis spectrum illnesses exhibit significant clinical and cognitive heterogeneity in symptoms, illness trajectory, and treatment response.^1^ This heterogeneity poses challenges for understanding the underlying pathophysiology and developing more efficacious individualized treatment strategies. Computational psychiatry has emerged as a promising framework for parsing interindividual variability beyond observed behavior.^2^ Modeling of trial-by-trial behavior can reveal distinct latent strategies used in goal-directed decision-making and offer insights into individualized cognitive profiles.^3^

In this context, we use the term “microcognitive processes” to refer descriptively to latent, trial-level computational processes, such as learning rates and decision noise, typically captured by computational model parameters. These fine-grained processes are not directly observable in standard cognitive testing but may jointly give rise to higher-level cognitive constructs typically measured in clinical settings. Computational modeling of such microcognitive decision-making mechanisms may illuminate why goal-directed behavior is affected in different ways across people with psychosis, potentially leading to new avenues for targeted interventions.

Adaptive decision making requires balancing the decision to sustain known rewarding actions against the decision to try new options, often referred to as “the exploration-exploitation tradeoff.” Disruptions in exploration-exploitation behavior have been frequently reported in psychosis spectrum illness, most notably excessive choice switching and less persistent exploitation.^4–14^ Some studies have implicated reward learning deficits such that patients failed to learn which choices were rewarding, leading to excessive switching.^11–13^ Other studies point to elevated decision noise or random exploration, meaning that patients’ choices were more variable and less driven by expected value.^15^ But the exact contributions made by each of these potential mechanisms remains unexplained.

More recent computational modeling approaches have identified latent microcognitive processes that contribute to alterations in exploration-exploitation tradeoff in psychosis.^15,16^ These parameters have revealed meaningful computational phenotypes with different underlying impairments. Examples include a schizophrenia subtype showing extreme ambiguity linked to poor cognitive functioning,^16^ and a schizophrenia subtype showing reward-based effort allocation disruptions linked to worse symptoms and functioning.^17^ These findings highlight key microcognitive processes that integrate reward and uncertainty for adaptive decision-making, demonstrating the potential for computational modeling to parse decision-making profiles within the psychosis spectrum.

Despite these computational advancements, prior value-based decision-making research has focused on reward learning under relatively stable reward contingencies, leaving gaps in understanding how people adapt under uncertainty. By modeling behavior on a dynamic restless bandit task, we examined how individuals transition between exploration and exploitation based on observed feedback and perceived volatility. We tested 75 participants with early psychosis (EP) and 68 neurotypical controls. Despite similar overall performance, EP exhibited excessive choice switching, which was driven by suboptimal transitions into exploration. These transitions, not explained by reward learning deficits, were linked to poorer real-world functioning.

Computational models identified two independent contributors to over-exploration, uncertainty sensitivity and decision noise, and revealed distinct EP subtypes linked to unique cognitive and clinical profiles. These findings extend prior work by identifying distinct microcognitive processes that independently disrupt adaptive decision-making and highlighting potential neurophysiological targets for personalized treatment.

## Methods

### Participants

Participants (15-45 years old) were recruited as part of a larger longitudinal neuroimaging study at the University of Minnesota through targeted online advertising and, for participants with Early Psychosis (EP), local psychiatry clinics. Demographics were matched across groups with the exception of gender: 75 EP (mean age 25.0 ± 5.1 years; 38.7% cis-woman, 29.3% cis-man, 32.0% other; 70.7% White) and 68 controls with no history of psychosis (mean age 26.5 ± 6.8 years; 54.4% cis-woman, 39.7% cis-man, 5.9% other; 72.1% White). A total of 143 participants (**Table S1**) completed the first session, and 124 participants (*n*=61 EP; *n*=63 controls) returned for the second session of the study approximately 3 weeks later to examine test-retest reliability. Behavioral data from both sessions were used in analyses; fMRI results will be reported elsewhere. Participants received compensation and bonus payments for task performance. Additional details in Supplement.

### Clinical and Self-report Measures

Participants completed clinical interviews and questionnaires assessing symptoms and functioning after consenting. The DSM-5-based Mini International Neuropsychiatric Interview was used to assess presence of diagnosis in EP and controls. Dimensional psychiatric symptom severity was measured using the Brief Psychiatric Rating Scale-24 Item Version^18^ across all participants. The Scales for the Assessment of Negative/Positive Symptoms^19,20^ provided psychosis-related symptom severity ratings in EP. Cognitive functioning was assessed via Test My Brain tasks^21^, capturing processing speed (digit symbol matching), executive function (matrix reasoning), verbal memory (verbal paired associates), social cognition (emotion recognition), and estimated IQ (based on verbal memory). Test My Brain is a research platform hosted by the Many Brains Project nonprofit organization, and a digital open science tool that provides brief web-based cognitive assessments that are automatically scored using normed data. Additional details in Supplement.

### Behavioral Task: three-armed restless Bandit Task

Participants performed a three-armed restless bandit task^22^, a dynamic decision-making paradigm involving stochastic rewards (**Figure 1A**). In each trial, three Gabor patch images were presented, each associated with a hidden probability of reward (+1 or 0), which drifted independently and randomly over time (**Figure 1B**). Each arm’s reward probability had a 10% chance of increasing or decreasing by 0.1 per trial. Participants were incentivized to maximize points, balancing exploration and exploitation using feedback. This design encouraged individual variability in decision-making strategies. A 200-trial version was administered during 3T fMRI, followed ∼3 weeks later by a 300-trial version with concurrent fMRI and EEG, each preceded by 25 practice trials (see Supplement).

**Figure 1.**
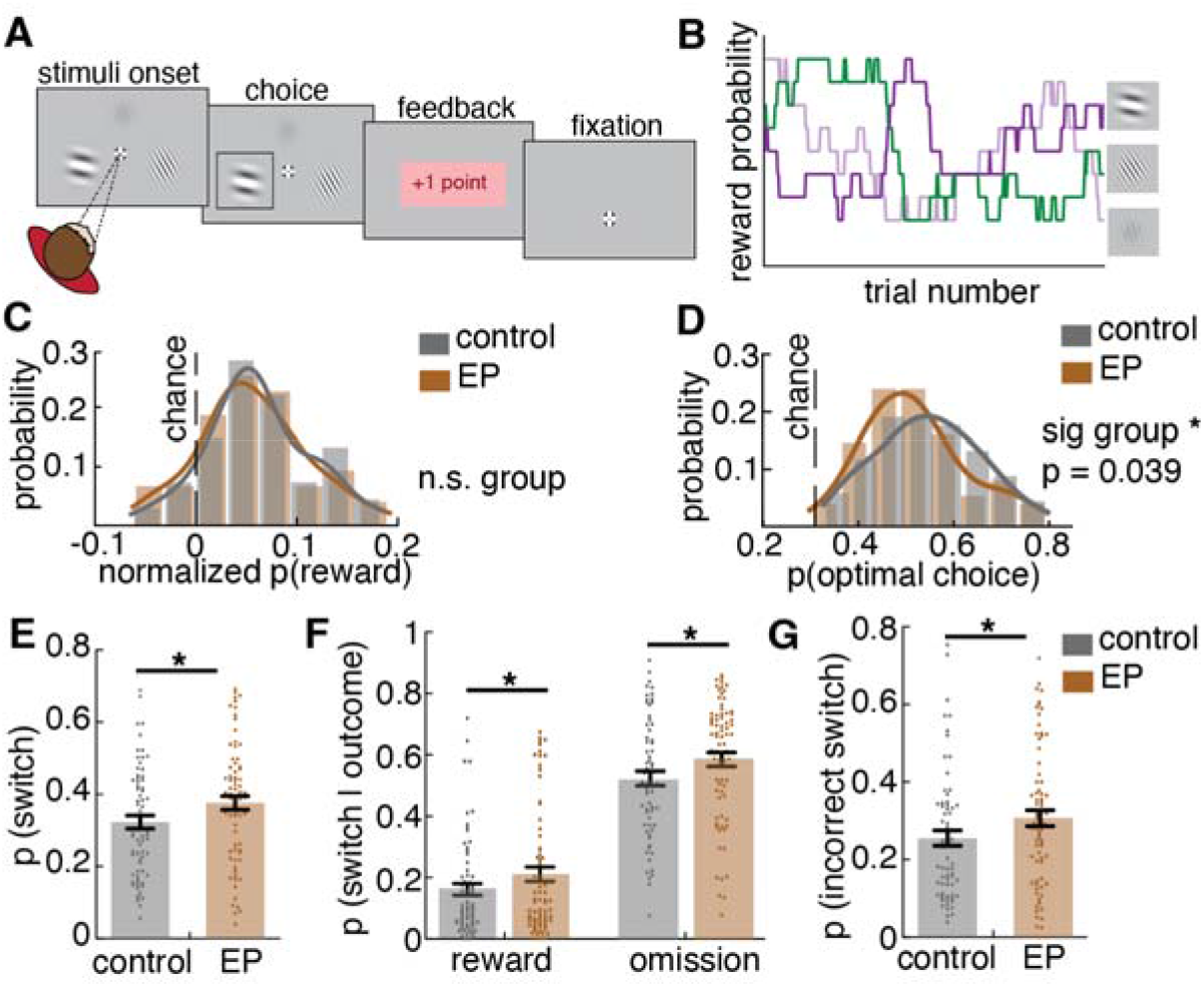
Participants with Early Psychosis were over switching, regardless of the outcome and the most optimal choice in the restless Bandit Task. **A**) task schematic. A three-armed restless Bandit task, in which participants chose from one of three gabor patch stimuli and received feedback of reward (+1 point or +0 points). Reward was probabilistic. Participants completed the restless Bandit task at two sessions, approximately three weeks apart. Data from both timepoints were used in analyses. **B**) An example of dynamic reward contingency where the probability of reward associated with each gabor stimulus changed slowly, independently, and randomly over time, **C**) Average reward acquisition normalized by chance level. **D**) Average probability of choosing the optimal choice (highest payoff choice). EPs were less likely to choose the optimal choice. **E**) Average probability of switching to a different choice, **F**) Average probability of switch given a rewarded or reward omission outcome in the previous trial (i.e. win switch and lose switch). **G**) Average probability of incorrect switch (switching away from an optimal choice). Since there were no prominent session effects, only session 1 data is shown here for clarity. Session 2 results can be found in Figure S1. * indicates p < 0.05. Graphs depict mean ± SEM across participants.

### Statistical techniques

Data were analyzed using custom PYTHON/R scripts, Prism 9 and SPSS. Generalized linear mixed models (GLMMs), ANOVAs, and t-tests assessed group effects across sessions. Group (EP, control) and session were modeled as fixed effects, participant ID as a random effect, and group x session interactions captured differential change over time. Since results were qualitatively similar across sessions, only session 1 results are shown in main figures; session 2 results are reported in Supplement. Gender and chance-level reward variability were included as covariates due to group differences and slight session-to-session random chance drift. Spearman correlations examined associations between task parameters and clinical features using session 1 data, as clinical assessments preceded this session and were not repeated.

Statistical significance was assessed at □ = .05, unless otherwise noted.

### Hidden Markov Model (HMM)

We fit a Hidden Markov Model (HMM) to infer latent states (explore/exploit) from participant choice sequences.^22,23^ Fitting a mixture model of exponential distribution revealed two latent strategies. Details on the mixture model and the HMM model can be found in Supplement. The Hidden Markov Model used to infer latent exploration states has also undergone parameter recovery validation in the same task ^24^, supporting the interpretability of the modeling framework used here. We fit the HMM model to each participant’s choice sequence using the Baum Welch algorithm,^25^ extract individualized transition dynamics of explore-exploit states, and label each choice based on its most probable state. Further analysis of the transition matrix revealed the dynamics of choice behaviors (See Supplement).

### Bayesian learner model (Kalman filter) and Reinforcement Learning (RL) models

We fit Bayesian learner models based on a Kalman filter framework,^26,27^ which adapts learning rate based on estimated uncertainty. In these models, action selection depends on estimated value, estimated uncertainty, and value-independent choice persistence, with a Softmax inverse temperature parameter controlling decision noise. The primary model includes three free parameters: uncertainty sensitivity φ, perseveration weight ω, and inverse temperature β. We also evaluated an extended Bayesian learner model incorporating a choice kernel to capture longer-range choice repetition effects.

For comparison, we fitted the following reinforcement learning (RL) models: 1) **RL+perseveration**: incorporates a first-order perseveration term, 2) **RL+choice kernel**: a basic delta-rule RL model with an additional choice kernel updating rule which captures the value-independent choice persistence, 3) **asymmetric learning + choice kernel** model: incorporates an additional asymmetric learning scalar parameter that scales the learning rate on the trials where there is no reward, allowing for different learning rate for positive and negative prediction error, 4) **Dual-state RL**: assumes two learning rates for HMM-inferred explore and exploit state. Details on the Bayesian learner models and RL models can be found in Supplement. To assess the necessity of uncertainty sensitivity (φ) and perseveration (ω) parameters beyond Bayesian belief updating, we performed nested model ablations within the Kalman filter framework. Reduced models excluded φ, ω, or both, while preserving identical learning dynamics. Models were compared using negative log-likelihood and BIC to account for differences in model complexity.

Model parameters were estimated separately for each participant using maximum likelihood estimation (MLE) by minimizing the negative log-likelihood of observed choices. Optimization was performed in Python using SciPy’s scipy.optimize.minimize function with the L-BFGS-B algorithm. Parameters were constrained to plausible ranges (inverse temperature β ∈ [0.1, 10], uncertainty sensitivity φ ∈ [0.01, 10], perseveration ω ∈ [0, 1]). To reduce sensitivity to local minima, optimization was repeated from 30 random initializations for each participant, retaining the best-fitting solution. Model comparison was performed using Akaike Information Criterion (AIC) and Bayesian Information Criterion (BIC), which account for both goodness of fit and model complexity.

### Hierarchical Clustering

Hierarchical clustering with Ward’s linkage method^28^ was applied to two bandit task-based computational parameters and three TestMyBrain measures from all participants across sessions. Features were selected to capture informative dimensions of decision-making relevant to the task, integrating computational parameters that shaped exploration behavior and showed group differences (uncertainty sensitivity and decision noise) with traditional cognitive domains that differed between groups and are closely related to learning and cognitive control (verbal memory, executive function, and processing speed; social cognition was excluded because the task does not involve social decision-making). Perseveration was not included because it did not differ between groups or contribute to over-exploration and therefore may not represent a major axis of informative variability in this dataset. A three-cluster solution was selected based on internal validity indices (**Figure S9**). The average silhouette coefficient peaked at *k* = 3, and the largest improvement across metrics occurred between two and three clusters, whereas additional clusters yielded only gradual, monotonic changes in Calinski-Harabasz and Davies-Bouldin indices alongside reduced silhouette values. Together, these findings indicate that the three-cluster model provided the most parsimonious representation of the data without evidence of meaningful additional separation at higher values of *k*. Clusters were characterized using the original computational and cognitive measures (**Figure 4C**).

## Results

### Task performance was comparable between participants with early psychosis (EP) and controls, but EP switched choices more often than controls regardless of the outcome

We examined performance based on total rewards (relative to chance) and frequency of optimal choices (defined as selecting the option with the highest underlying (unobservable) reward probability on a given trial; when multiple options shared the same highest reward probability, all such options were considered optimal). Both groups performed significantly above chance in session 1 (**Figure 1C;** one sample t-test, control: *t*(67) = 9.86, *p*<.0001, EP: *t*(74) = 9.0, *p*<.0001) and session 2 (**Figure S1**: control: *t*(62) = 12.41, *p*<.0001, EP: *t*(60) = 8.23, *p*<.0001), with no significant group differences in overall rewards (*F*(1,141)=2.18, *p*=.142).

However, participants with EP made fewer optimal choices (**Figure 1D**; *F*(1,141)=4.32, *p*=.039). To probe decision-making strategies, we analyzed stay/switch probabilities as an index of how participants evaluated environmental stability. Despite similar reward outcomes, participants with EP showed increased switching compared to controls (**Figure 1E**; *F*(1,141)=7.67, *p*=.006), including after rewards (win-switch; **Figure 1F**, *F*(1,141)=3.94, *p*=.049), omissions (**Figure 1F**, *F*(1,141)=7.53, *p*=.007), and away from optimal options (“incorrect switch”; **Figure 1G**, *F*(1,141)=5.17, *p*=.024). This suggests participants with EP engaged in more switching behavior regardless of the outcome, reducing optimal choices over time, consistent with prior literature.^4–14,29^

### Participants with EP explored more due to instability in exploration-exploitation strategy

To understand what drives increased switching, we examined explore–exploit strategies. Mixture modeling confirmed both groups used fast-switching (exploration) and slow-switching (exploitation) modes (**Figure S2A-B**; see **Supplement**). Using a Hidden Markov Model (HMM), we identified latent strategy states from individual choice sequences (**Figure 2A**), with exploration marked by distributed choices and exploitation by repeated selections (**Figure 2B**). Participants with EP explored more than controls (**Figure 2C**, *F*(1,141)=7.08, *p*=.009), consistent across sessions (**Figure S2C**). Within HMM-defined states, EP showed increased switching during exploitation (**Figure 2D**, *F*(1,138) = 5.53, *p* = .020), but not during exploration (*F*(1,141) = 0.03, *p*=.871), indicating unstable exploitation rather than generalized over-switching.

**Figure 2.**
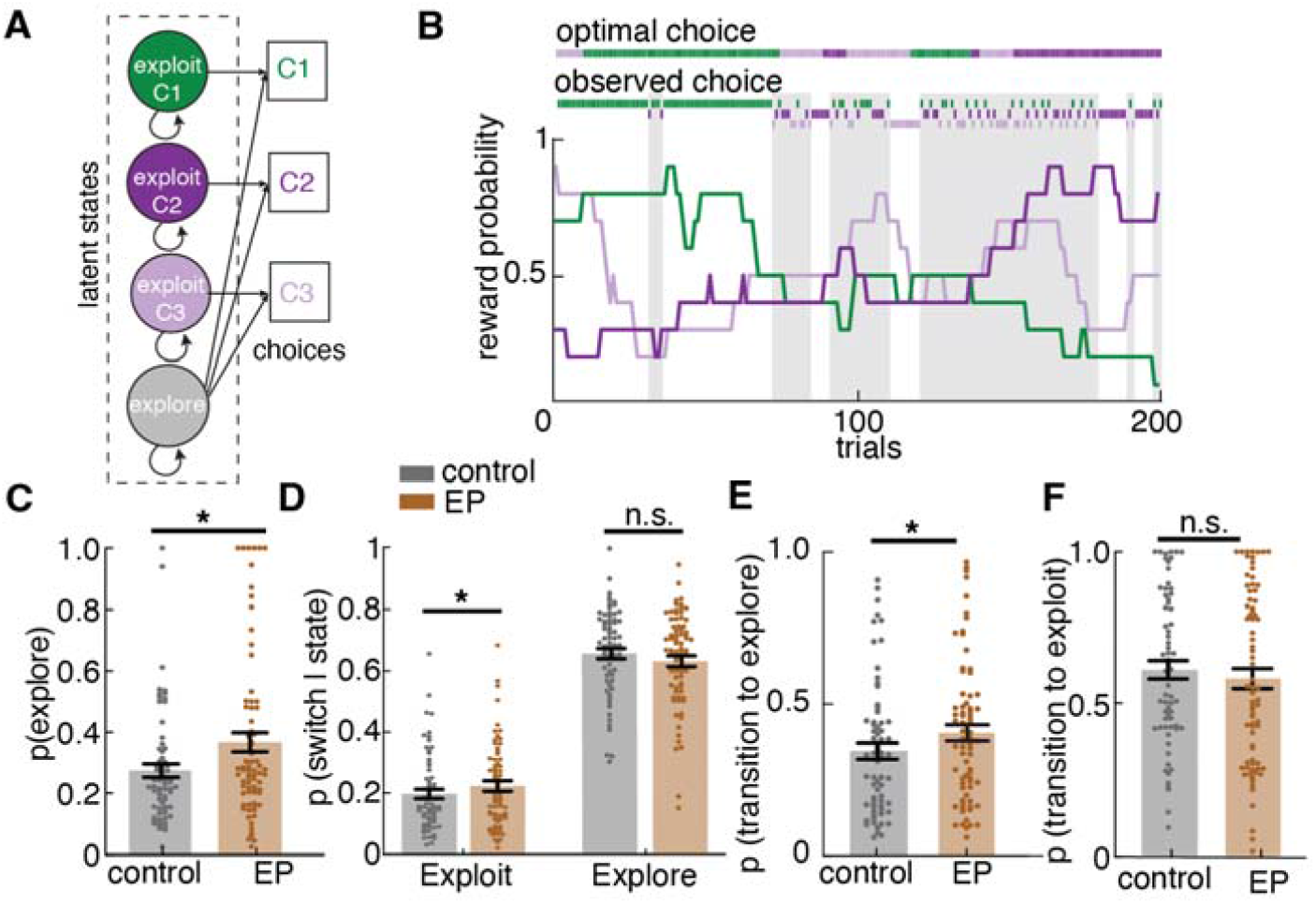
Participants with Early Psychosis over-explored due to a higher probability of transitioning into the explore strategy state, compared to controls. **A**) Hidden Markov model (HMM) model structure. The HMM models exploration and exploitation as latent strategy states underlying observed choices. This model includes an exploitation state for each choice, and an exploration state. **B**) Reward probabilities (colored lines), observed choices (lower dots), optimal choices defined by the highest payoff (upper dots), and HMM-labeled exploratory choices (shaded area) in an example session of 200 trials. **C**) Participants with Early Psychosis (EP) had increased exploration. **D**) Participants with EP had higher probability of switching during exploit strategy state. No group difference in the probability of switching during explore strategy state. **E**) EP had a higher probability of transitioning to explore strategy state (fitted transition matrix parameter), resulting in over exploration in EP. **F**) No group difference in the probability of transitioning to exploit strategy state. Since there were no session effects, only session 1 data is shown here for clarity. Session 2 results can be found in Figure S2. * indicates p < 0.05. Graphs depict mean ± SEM across participants.

Analysis of state transitions revealed participants with EP were more likely to switch from exploit to explore (**Figure 2E**, *F*(1,141)=8.32, *p*=.005), particularly after receiving a reward (*F*(1,141)=8.72, *p*=.004), indicating instability in the reward-driven exploit strategy state, not fully explained by loss avoidance (**Figure S2F**). However, transitions from explore to exploit were intact (**Figure 2F**), indicating preserved reward learning. These results suggest that the over-exploration in EP was not simply explained by reward learning deficits. So, what drove participants with EP to transition out of the exploit strategy state and start exploring?

### Over-exploration in EP was driven by increased uncertainty sensitivity and decision noise

To identify the computational mechanisms contributing to exploration behavior, we compared several classes of learning models, including Bayesian learner models based on a Kalman filter and reinforcement learning (RL) models with fixed learning rates (**Figure 3A**). The Bayesian learner models estimate both expected value and uncertainty for each option and adapt the effective learning rate based on uncertainty (**Figure 3B**). RL models, in contrast, update value estimates using fixed learning rates based on reward prediction errors. We evaluated multiple RL variants commonly used in bandit tasks, including models with a choice kernel capturing value-independent choice persistence, asymmetric learning rates for positive and negative prediction errors, and a dual-state RL model with separate learning rates during exploration and exploitation states. We also compared alternative formulations of choice persistence using either a first-order perseveration term or a choice kernel that captures longer-range choice history effects. Model comparison using information criteria (ΔBIC relative to the best model) favored a Bayesian learner model with first-order perseveration in Early Psychosis (**Figure 3A**). While model fits varied across groups, this was the best fitting model for our clinical group representing the population of interest. This model includes three parameters: uncertainty sensitivity (φ) which scales the influence of estimated uncertainty on choice; inverse temperature (β) which governs choice stochasticity, and perseveration parameter (ω) which captures value-independent choice stickiness. Full model specifications and comparison details are described in the supplement. Simulations confirmed that increases in φ and decreases in β promote exploration (**Figure S3A**), as well as win-switch behaviors (**Figure S3B**). **Figure 3C** illustrates an example model fit, showing trial-wise value and uncertainty estimates. We tested whether uncertainty sensitivity (φ) and perseveration (ω) add explanatory value beyond standard Bayesian learning by systematically removing them from a Kalman filter model and comparing model performance. The full model consistently outperformed with a lower negative log-likelihood and lower BIC than reduced models, in both control and early psychosis groups (**Figure S4A**). These improvements remained substantial despite penalization for additional parameters, indicating that the winning model’s advantage is not solely attributable to the Kalman learning rule. Parameter recovery analyses for the winning Bayesian Learner model demonstrated strong recoverability of key parameters (**Fig. S4B; Table S2**). This model builds on the SMEP framework introduced by Peters and colleagues, which has been previously validated through extensive parameter and model recovery analyses in related restless bandit tasks ^30^.

**Figure 3.**
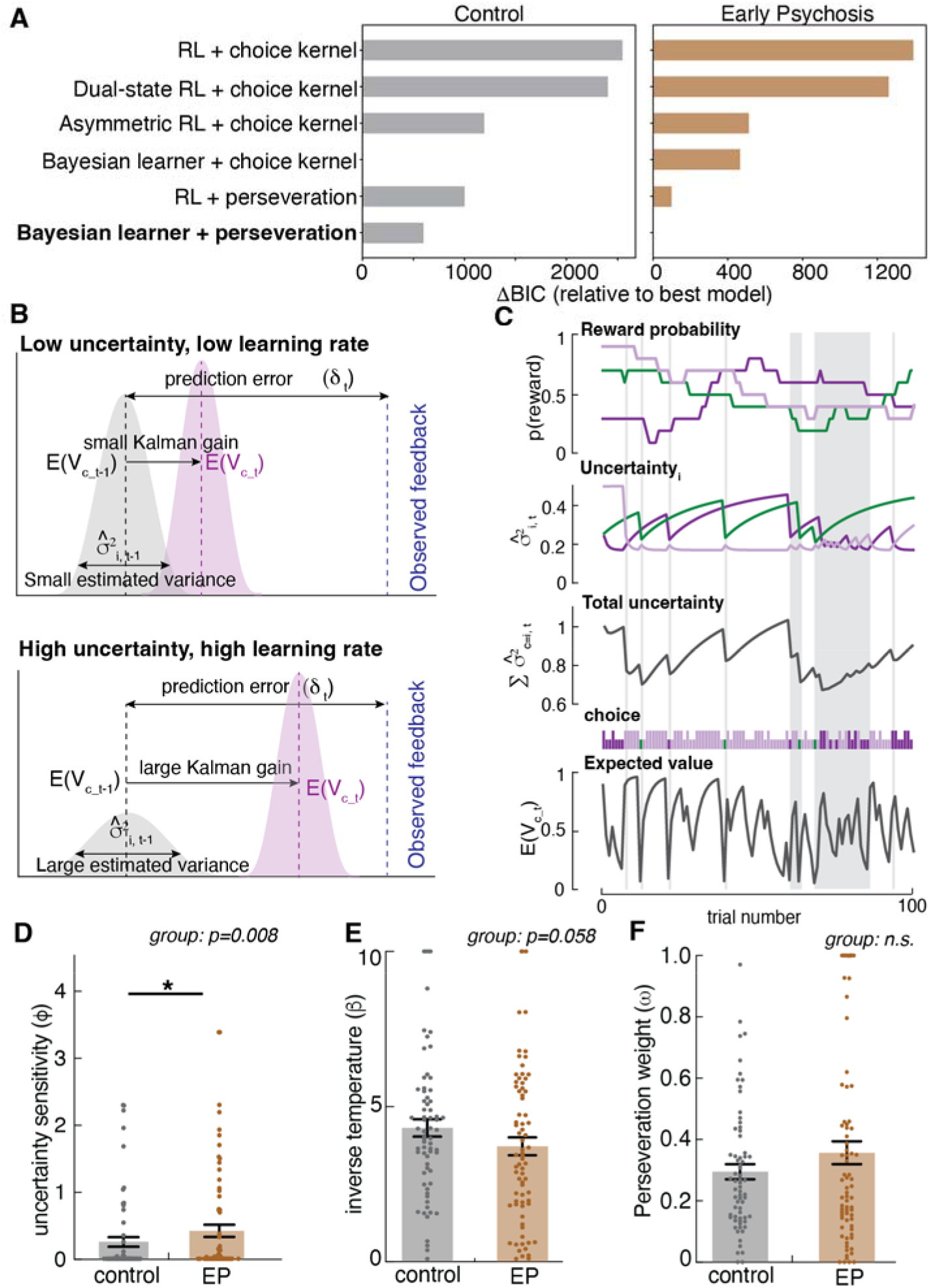
Participants with Early Psychosis had higher decision noise and uncertainty sensitivity inferred by a Bayesian learner model, and both processes contributed to over exploration in EP. **A)** Model comparison results for control and early psychosis groups using ΔBIC (relative to best-fitting model within each group). Lower values indicate better model fit after penalizing for model complexity. The Bayesian learner model with first-order perseveration provided the best overall fit under BIC in the Early Psychosis group. **B**) An illustration of the adaptive learning rate (Kalman gain) under low and high uncertainty. When uncertainty, captured by the variance of estimated value distribution, is low, the most recent feedback is weighted less and the learned value from reward history is weighted more, less of the prediction error is being updated, the Kalman gain (learning rate) is smaller. When uncertainty is high, the most recent feedback is weighted more, the Kalman gain (learning rate) is larger, which means more of the prediction error is updated. In this model, when uncertainty is high, the model increases learning rate, promoting exploration; when uncertainty is low, the model relies more on learned value, promoting exploitation. **C**) An example of Bayesian Learner model fit to participant data (example of 100 trials) and trial-wise value estimation as well as variance (uncertainty) estimates. The uncertainty level towards an unselected choice gradually increases over time and reduces when being selected. The choice selection is dependent on both expected value and uncertainty of three choices, with some random noise. **D**) Uncertainty sensitivity (φ) fitted to participant choice sequences in Session 1. EPs had higher uncertainty sensitivity compared to controls. **E**) Inverse temperature (β) fitted to participants in Session 1. EPs had a trend of lower inverse temperature (higher decision noise) compared to controls. **F**) Perseveration weight (ω) fitted to participants in Session 1. There was no group difference in perseveration weight. Since there were no session effects, only session 1 data is shown here for clarity. Session 2 results can be found in Figure S5. * indicates *p* < .05. Graphs depict mean ± SEM across participants.

For ease of interpretation, we refer to φ as uncertainty sensitivity and to β^-1^ as decision noise throughout, while emphasizing that these labels are descriptive interpretations of the computational model parameters. Outlier detection identified 4 uncertainty sensitivity parameter data points with values that were winsorized for analyses. Participants with EP showed significantly higher uncertainty sensitivity than controls across sessions (**Figure 3D**; *F*(1,141)=7.27, *p*=.008). They also trended toward greater decision noise than controls across sessions (**Figure 3E**; *F*(1,141)=3.64, *p*=.058), indicating less consistent reliance on learned value. No group difference was found in perseveration (**Figure 3F**). Session 2 plots can be found in Figure S5. Thus, increased exploration in EP reflects a stronger drive to reduce uncertainty and noisier decision-making.

### Exploratory analysis: Individual differences in exploratory strategy were linked to distinct computational subtypes

To determine whether variability in computational and cognitive measures reflected a uniform shift associated with diagnosis or organized into latent subgroups independent of diagnostic status, we conducted hierarchical clustering using Ward’s linkage ^28^ on a five-dimensional behavioral feature space (uncertainty sensitivity, inverse temperature, processing speed accuracy, verbal memory accuracy, and executive function accuracy) (**Figure 4A, Figure S6**). Clusters were derived without reference to diagnosis, and the distribution of diagnostic groups across clusters was examined (**Figure 4B**). This approach allowed us to identify three distinct and potentially clinically meaningful computational subtypes among both controls and EPs. These subtypes were largely stable over time, with 86% retention in the same subtype across sessions. Although both groups expressed all subtypes, their distributions differed: controls were overrepresented in Subtype 1, while EP participants were overrepresented in Subtypes 2 and 3 (**Figure 4B**). Subtypes were profiled by mean levels of uncertainty sensitivity, decision noise, processing speed, verbal memory, and executive function (**Figure 4C**). For visualization in the radar plot, the more peripheral parameters reflect better cognitive performance and lower decision noise, whereas closer-to-center parameters indicate more severe impairments. Each cluster characterized a distinct computational subtype. For clarity, we refer to the three clusters as “Subtypes 1-3” throughout. Subtype 1 was largely normative, with low uncertainty sensitivity, low noise and strong cognition. Subtype 2 showed high uncertainty sensitivity, slow processing, and poor memory but intact executive function. Subtype 3 was marked by high decision noise and broad cognitive deficits, consistent with prior schizophrenia findings.^16^

**Figure 4.**
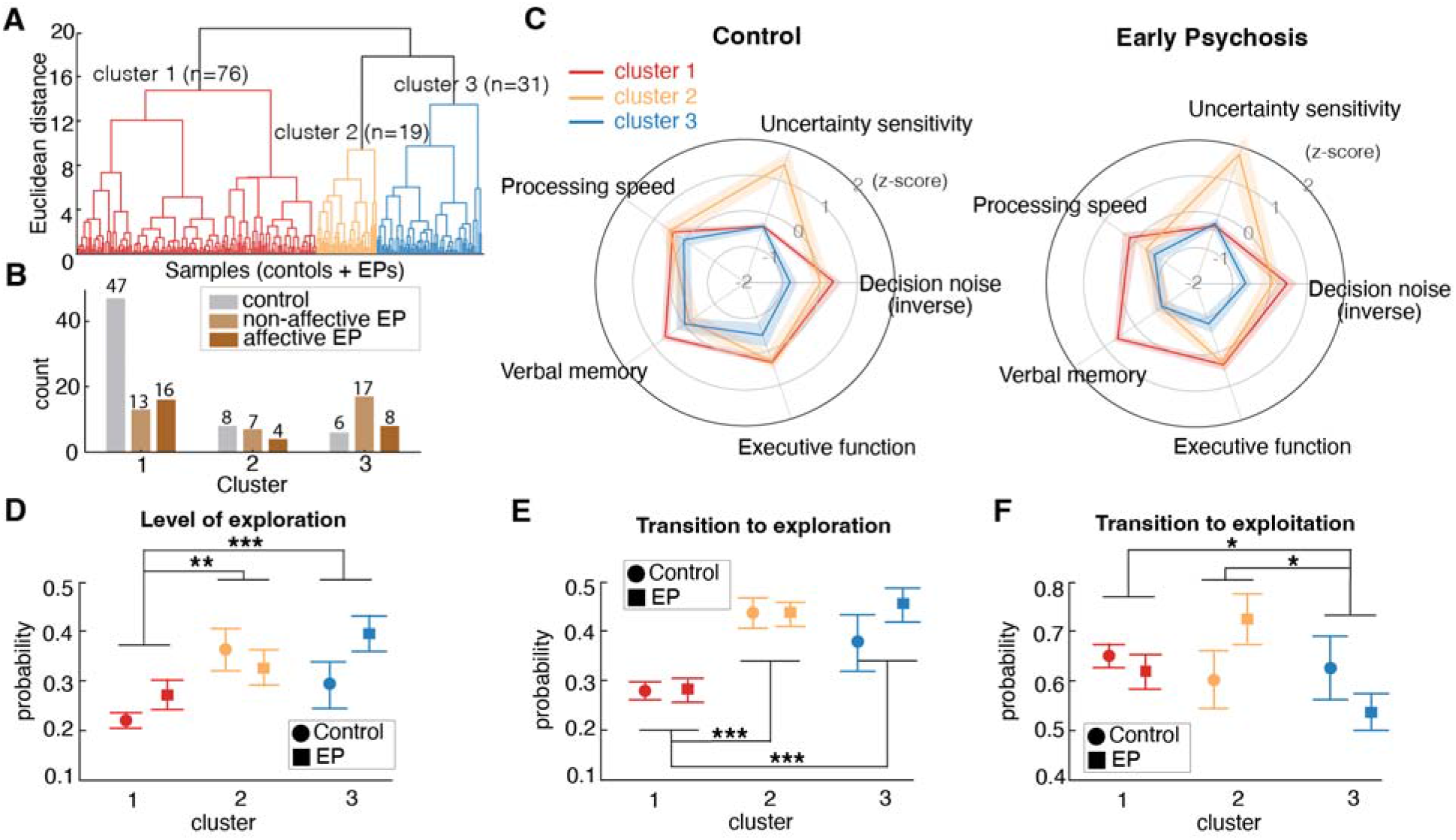
Task strategy revealed three computational subtypes that are associated with distinct cognitive profiles and exploration-exploitation tradeoff. **A**) Hierarchical clustering on the computational and cognitive processing measures from session 1 identified three clusters across controls and EPs: 1-red (n=76), 2-orange (n = 19), and 3-blue (n = 31). **B)** Distribution of controls, affective and non-affective EPs in each cluster. Controls predominantly occupied cluster 1 while EPs were over-represented in clusters 3. **C)** Radar plot depicting cluster-specific computational and cognitive profiles in controls and EPs. Each cluster represents a distinct cognitive subtype. Notably, cluster patterns were similar across controls and EPs but more impaired in EPs. For visualization, decision noise were inverted. The more peripheral a cluster, the better cognitive performance and computational stability; the closer to the center, the more impaired the profile. Subtype 1 is characterized by low uncertainty sensitivity, low decision noise and strong performance across cognitive measures. Subtype 2 exhibits high uncertainty sensitivity but retains relatively intact executive function compared to cluster 3. Subtype 3 shows high decision noise and the most pronounced cognitive impairments across cognitive domains. **D)** Both Subtype 2 and Subtype 3 have the higher level of exploration than Subtype 1. **E)** Subtypes 2 and 3 have the higher probability of transition to exploration than Subtype 1. **F)** Subtype 3 has the lowest probability of transitioning to exploitation. Graphs in D-F depict mean ± SEM across subjects, statistics are reported in Table S3. Significant feature loadings are indicated by * *p*<.05, ** *p*<.01, *** *p*<.001.

A two-way ANOVA revealed a significant main effect of Subtype on exploration strategy use (all *p* values <.05) (**Table S3**). Post hoc pairwise comparisons were conducted using false discovery rate (FDR) correction to control for multiple comparisons. Subtype 1 explored the least, compared to subtype 2 (adjusted *p* =.002) and subtype 3 (adjusted *p*<.001) (**Figure 4D**). Both Subtypes 2 and 3 had elevated transitions to exploration compared to Subtype 1 (1 vs. 2: adjusted *p*<.001, 1 vs. 3: adjusted *p*<.001) (**Figure 4E**). Only Subtype 3 showed reduced transitions to exploitation compared to Subtype 1 (adjusted *p*=.048 and subtype 2 (adjusted *p*=.048) (**Figure 4F**). This transition dynamic is indicative of reward learning deficits because low probability of transition into exploitation suggests difficulty in learning about the highest valued choice to exploit. These findings suggest that although both Subtypes 2 and 3 initiated explorations prematurely, only Subtype 3 exhibited difficulty sustaining exploitation of high-value options, consistent with impaired reward learning.

### Exploration and task strategy on the restless Bandit task was associated with clinical features in individuals with Early Psychosis

Task strategy remained stable across two sessions in all subjects, with moderate test-retest reliability (see Intraclass Correlation Coefficients in **Figure S7**). To assess clinical relevance of task strategy within EP, we correlated three task variables (transitions to exploration, decision noise, uncertainty sensitivity) with six clinical measures (negative/ positive/ disorganized/ manic/ depressed symptoms, role functioning) using False Discovery Rate (FDR) corrected Spearman correlations (**Table S4**). Greater transition to exploration was associated with poorer real-world functioning^31,32^ (*n*=74, *rho* = -.40, FDR *p* = .014; **Figure 5A**) and higher decision noise (*n*=75, *rho* = -.77, FDR *p* < .001). Higher decision noise was associated with negative symptom severity (*n* = 74, *rho* = -.37, *p* = .045, **Figure 5B**) and showed a trend association with poorer real-world role functioning (*n* = 74, *rho* = .36, *p* = .055). We also compared exploration strategy across controls (*n*=68), non-affective EP (*n*=46), and affective EP (*n*=20). Exploration differed by group (*F*(2,140)=3.53, *p*=.032, **Figure 5C**), with trend-level elevations in both non-affective (*p*=.062) and affective (*p*=.099) EP subgroups. Transitions to exploration also differed (*F*(2,140)=4.30, *p*=.015, **Figure 5D**), driven by significantly higher rates in non-affective EP vs. controls (*p*=.019). These findings suggest that elevated exploration in EP is not solely attributable to mood-related pathology. Additional individual difference analyses in Supplement (**Figure S8**).

**Figure 5.**
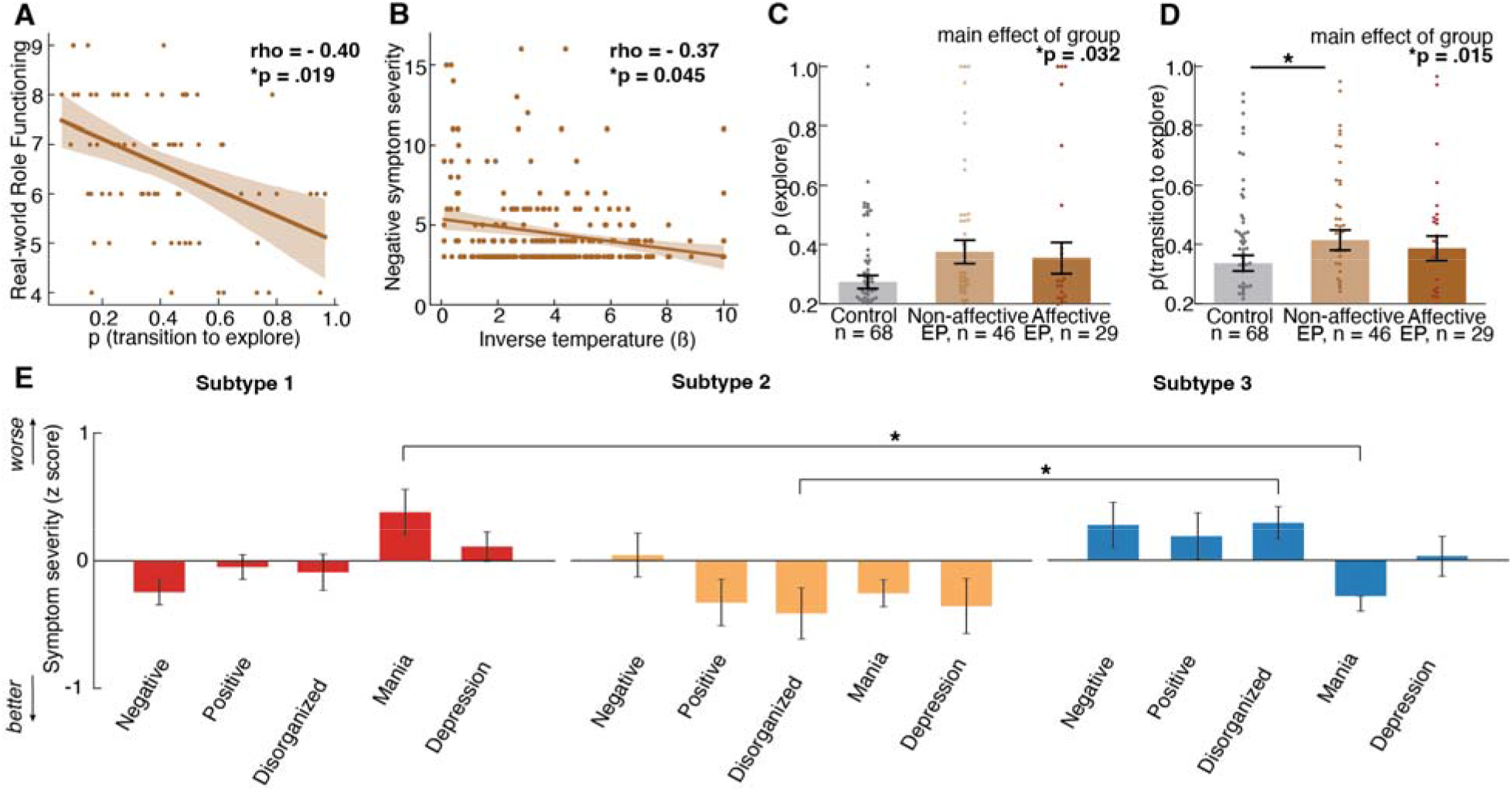
Task strategy was associated with symptom severity and real-world functioning among 74 participants with Early Psychosis (EP) at session 1. Symptom severity was measured with the Brief Psychiatric Rating Scale. **A**) Higher probability of transitioning to explore strategy state (fitted HMM transition matrix parameter) correlated with lower real-world functioning. **B**) Lower Bayesian model inverse temperature, indicating higher decision noise, correlated with more severe negative symptom severity. **C**) EP with non-affective psychosis (n=46) and EP with affective psychosis (n=29) had higher probability of exploration compared to controls (n=68). **D)** EP with non-affective psychosis had higher probability of transitioning into exploration strategy state, suggesting altered explore/exploit transition dynamics that is most pronounced in non-affective psychosis. **E)** Symptom severity was measured with the Brief Psychiatric Rating Scale (BPRS), including negative symptom, positive symptom, disorganized symptom, mania and depression symptom subscales. Average symptom severity score was calculated and standardized for each of the three subtypes: subtype 1 - normative, subtype 2 - uncertainty-sensitive, subtype 3 - decision noise with cognitive impairments. * indicates p < 0.05, ** p<0.01, *** p <0.001.

### Subtypes showed unique clinical correlates among Early Psychosis

Within the EP group, computational subtypes differed in disorganized (F(2,46)=3.88, *p*=.028) and manic symptom severity (*F*(2,45)=3.47, *p*=.040); **Figure 5E**). Subtype 3 (decision-noise subtype) had greater disorganized symptom severity than Subtype 2 (uncertainty-sensitive subtype, FDR corrected-*p*=.042). Subtype 1 (normative subtype) had greater manic symptom severity than Subtype 3 (FDR corrected-*p*=.030). Subtypes also varied in clinical history: Subtype 2 (uncertainty-sensitive subtype) had a higher rate of multiple lifetime psychiatric hospitalizations (*□* ^2^(2)=7.07, *p*=.029; adjusted Pearson residual = 2.60). The distribution of primary diagnoses varied significantly across subtypes (*□* ^2^(12)=25.25, *p*=.014. Subtype 1 showed a higher proportion of bipolar disorder with psychotic features (44.6%) compared with Subtype 2 (27.3%) and Subtype 3 (14.0%). Standardized residuals suggested relative overrepresentation of bipolar disorder in Cluster 1 (z=2.05) and underrepresentation in Cluster 3 (z=-2.04), though cell-wise tests did not survive correction due to small counts in some diagnosis cells. Subtypes did not differ by age (*p*=.771), illness duration (*p*=.394), or antipsychotic use (*p*=.399).

## Discussion

In this study, we applied computational modeling to characterize how individuals with early psychosis (EP) and controls adaptively navigate the tradeoff between exploiting known rewarding choices and exploring for information. Although overall reward performance was similar, EP were less likely to persist with the optimal choice due to a higher tendency to switch choices, a pattern observed across paradigms in EP but potentially arising from different computational mechanisms ^10^. While often attributed to impaired reward learning,^6,12–14^ our findings point instead to instability in goal-directed exploit strategy: EP were more likely to leave a favorable option and transition into exploration prematurely. Crucially, once a rewarding choice was identified, EP and controls showed similar tendencies to begin exploiting, indicating that the over-exploration in EP stemmed from difficulty sustaining, not initiating, value-based exploitation. This raises the question: why did participants with EP exit a favorable strategy too early?

### The role of uncertainty sensitivity

Our Bayesian learner modeling identified uncertainty sensitivity as a key driver of the suboptimal shift from exploitation to exploration in EP. While decision noise also contributed, uncertainty sensitivity emerged as a distinct mechanism shaping how estimated uncertainty influenced choice behavior. This aligns with theories of psychosis as a disorder of aberrant inference and altered representations of uncertainty.^6,33–35^ Individuals with psychosis have been reported to overestimate environmental volatility, leading to excessive choice switching.^6,36–38^ Cole et al. (2020) found clinical high-risk individuals overestimated volatility and showed abnormal prefrontal and insular responses to unexpected outcomes but blunted responses to belief updates, suggesting neural markers of uncertainty sensitivity may precede illness onset.^38^ However, interpretations of increased switching are inherently model dependent. Increased switching can arise from multiple computational mechanisms, including heightened uncertainty sensitivity (i.e. stronger uncertainty-guided or directed exploration) ^39–41^, increased decision noise ^16,41,42^, or greater inferred environmental volatility ^6,38^. Consistent with this, our results highlight substantial heterogeneity within early psychosis, such that increased exploration and switching behavior can arise from distinct computational mechanisms, including elevated decision noise or heightened uncertainty sensitivity. Behavioral responses to uncertainty can also vary: some over-explore to reduce ambiguity,^6,36–38^ while others exhibit strong ambiguity aversion.^16^ Because the present task contains no explicit cost for exploration, it was optimized to estimate uncertainty-bonus-driven exploration rather than to dissociate uncertainty seeking from ambiguity aversion ^16^. Behavioral responses to uncertainty may reflect both task context ^43^ and commonly co-occurring mood and anxiety symptoms linked to heightened uncertainty sensitivity.^44–48^ However, we found no evidence that affective symptoms explained decision-making instability in EP. Neural evidence implicates the salience network, particularly the anterior insula, in processing uncertainty,^49–51^ with hyperactivation observed in psychosis,^52,53^ anxiety.^54–56^

### The role of decision noise

Elevated decision noise also contributed to over-exploration in EP. This aligns with prior findings linking increased choice randomness (often modeled as lower inverse temperature) to frequent switching behavior.^57^ A recent meta-analysis identified heightened decision noise as a key mechanism underlying the increased choice switching in psychosis.^43^ Decision noise likely reflects neuromodulatory disruptions, particularly in dopamine and norepinephrine systems.^41,42,58^ Systemic dopamine manipulations bidirectionally modulated decision noise in mice.^42^ Noradrenergic tone predicted changes in exploration-exploitation tradeoff in healthy humans.^59^ Our findings suggest both uncertainty-driven (“directed”) and noise-driven (“random”) exploration contribute to excessive switching in EP but decision noise alone does not fully capture the complexity of disrupted explore-exploit dynamics in EP.

### Computational subtyping

Hierarchical clustering revealed three distinct computational subtypes in EP with unique explore/exploit dynamics and cognition profiles and were stable across sessions. This stratification parallels the three psychosis biotypes identified by the B-SNIP (Bipolar-Schizophrenia Network on Intermediate Phenotypes) consortium using biomarkers.^60,61^ Our decision noise subtype mirrors B-SNIP Biotype 1 (marked cognitive dysfunction, low neural response); our uncertainty-sensitive subtype aligns with Biotype 2 (hyperactive neural responses, moderate cognitive control deficits); and our normative subtype resembles Biotype 3 (near-normal cognitive and sensorimotor profiles). Consistent with B-SNIP findings, diagnostic distributions differed by subtype: bipolar disorder with psychosis was over-represented in the normative subtype, while schizophrenia was more prevalent in the other two. These findings highlight substantial cognitive and computational heterogeneity often missed by diagnostic classification, underscoring the need for precision psychiatry.^3^ The overlap with B-SNIP biotypes suggests that future cross-study validation may show that computational modeling of decision-making could offer a scalable alternative to biomarker panels. More importantly, these subtypes may guide mechanism-based treatments. Beyond characterization and subtype parallels with B-SNIP biotypes, computational phenotypes may hold promise for predicting treatment response. For example, Hauke et al. (2022) found that increased belief instability, which is a computational parameter reflecting rapid belief updating, predicted successful response to metacognitive training in individuals with psychotic disorders.^62^ Although the specific parameters and tasks differ between studies, this work supports the broader translational potential of computational stratification in identifying individuals who may benefit from targeted interventions. Overall, this computational subtyping approach offers a promising, accessible framework for individualized treatment in psychosis.

### Association with clinical features

Computational parameters demonstrated moderate test-retest reliability across sessions, supporting their use in linking behavior to clinical features. More severe negative symptoms and poorer real-world functioning were associated with increased suboptimal transition into exploration and high decision noise. Uncertainty sensitivity was not significantly correlated with symptom severity, potentially due to sample size limitations or an indirect mapping between symptom ratings and underlying cognitive processes.

Subtyping analysis revealed more nuanced findings than group-level associations, capturing individual differences better than symptom severity or diagnosis alone. The mood-predominant subtype (Subtype 1) showed normative learning and explore-exploit behavior, unlike the schizophrenia-spectrum subtypes (Subtypes 2 and 3), both marked by over-exploration. However, Subtypes 2 and 3 had different underlying computational contributors and clinical correlates - mainly, decision noise and poor reward learning map onto a subtype with elevated negative symptom severity whereas uncertainty sensitivity maps onto a subtype with elevated psychiatric hospitalization history, possibly reflecting (perceived) volatility that may shape how they balance explore-exploit.

### Reliability of computational measures

Reliability of computational measures has been an active area of investigation in recent years, with several studies reporting moderate to poor reliability for reinforcement-learning parameters and limited consistency across tasks.^63–66^ These findings suggest that computational parameters are often task- and model-dependent, reflecting specific cognitive processes engaged by a given paradigm rather than stable traits that generalize across all tasks. Consistent with this perspective, our task was optimized to quantify uncertainty-bonus-driven exploration. Nevertheless, within this paradigm we observed moderate-high test-retest reliability for several behavioral and computational measures, particularly inverse temperature, HMM-inferred exploration, and model-free learning metrics, despite participants completing different random reward walks across sessions. This suggests that the individual differences identified here reflect relatively stable decision-making strategies within the task context, despite unique reward contingencies in each session. This highlights the need for future studies to also evaluate psychometric properties of the computational measures, including retest reliability and linking clinical correlates, among others.

### Study limitations

Several limitations of the study should be acknowledged. First, gender distributions differed between groups, so gender was included as a covariate in analyses. Second, due to the randomized reward structure, EP experienced slightly lower reward availability in session 1; this was controlled for in analyses and did not meaningfully affect behavior (**Figure S1-2**). Third, the cross-sectional design limits evaluation of the long-term stability and predictive validity of computational measures. Lastly, future studies could incorporate broader self-report traits (e.g., uncertainty tolerance, anxiety) to deepen insights into mechanisms driving heterogeneity in decision-making in psychosis.

### Translational Potential

The restless bandit task offers strong translational and scalable potential by revealing interindividual variability in decision-making strategies with clinical relevance. Shared cognitive processes across species, this explore-exploit task is well-suited for both human clinical application and mechanistic animal experiments, linking neural, pharmacological, and genetic mechanisms to decision-making.^23,67,68^ Recent work shows humans, primates, and mice balance exploratory and exploitative strategies,^69^ supported by shared neural circuits, such as the anterior insula and frontoparietal network.^70^ By quantifying individual explore-exploit dynamics and translating mechanistic insights, the task supports the development of personalized “decision-making fingerprints” to inform targeted interventions.

### Conclusions

In this study, we found excessive choice switching in Early Psychosis on a value-based decision-making task was driven by suboptimal transitions into exploration. Uncertainty sensitivity and decision noise emerged as key cognitive processes underlying this over exploration. The study identified distinct computational subtypes with unique symptom profiles: a high decision-noise subtype with reward learning deficits and worse negative symptoms, a normative subtype with worse mood symptoms, and a novel uncertainty-sensitive subtype with higher hospitalization rates. The uncertainty-sensitive subtype was free of reward learning deficits and clinically undetectable in terms of symptom severity. These findings offer preliminary evidence supporting the value of computational modeling in precision psychiatry for early psychosis.

## Data Availability

All data produced in the present study are available upon reasonable request to the authors. All data will be made publicly available through the National Data Archive.

https://nda.nih.gov/

## Funding

This work was supported by the National Institute of Mental Health of the National Institutes of Health (1P50MH119569-01A1 to S.V. and A.D.R.).

## Acknowledgements

We would like to thank the participants who made this work possible. We would also like to thank our study staff who completed the data collection for this study as well as our collaborator Bryon Mueller who provided technical support and developed the neuroimaging protocol for task data collection.

## Notes

### Competing Interest Statement

The authors have declared no competing interest.

### Funding Statement

Research reported in this publication was supported by the National Institute of Mental Health of the National Institutes of Health under Award Number 1P50MH119569-01A1 (MPI Vinogradov, Redish). The content is solely the responsibility of the authors and does not necessarily represent the official views of the National Institutes of Health.

### Author Declarations

Ethics committee/IRB of the University of Minnesota gave ethical approval for this work

### Summary of Updates

Additional computational model testing (e.g., model comparison, parameter recovery) was completed.

