## Supplemental Materials for "Explore-exploit instability reveals computational decision-making heterogeneity in early psychosis"

Cathy S. Chen, Evan Knep, Veldon-James Laurie, Olivia Calvin, R. Becket Ebitz, Melissa Fisher, Michael-Paul Schallmo, Scott R. Sponheim, Matthew V. Chafee, Sarah R. Heilbronner, Nicola M. Grissom, A. David Redish, Angus W. MacDonald III, Sophia Vinogradov,

Caroline Demro

**Supplementary Methods.**

*Subjects:* Data from all enrolled participants who completed at least one baseline session were included for data analysis. Inclusion criteria for all participants were: 15-45 years old, English as a primary language, IQ≥70. Exclusion criteria for all participants were: current pregnancy; other MRI contraindications; severe substance use disorder; major neurological disorder; history of clinically significant head injury or significant cognitive training. For control participants, additional exclusion criteria were: current diagnosis or family history of psychotic, bipolar, or autism spectrum disorder. For participants with EP, additional inclusion criteria were: age 15-35 or 35-45 with onset of psychosis spectrum illness in the past 5 years. Written informed consent, or assent with the consent of a caregiver in the case of minors, was provided by all eligible participants and their capacity to provide informed consent was evaluated prior to study procedures using the University of California Brief Assessment of Capacity to Consent assessment tool^1^. All procedures were approved by the Institutional Review Board at University of Minnesota.

*Clinical Measures:* To assess suicidality, the Columbia Suicide Severity Rating Scale (C-SSRS) replaced the MINI suicidality module because it more succinctly assesses suicidal thoughts and behavior, reducing participant burden. Total negative and positive symptom severity was assessed with the SANS/SAPS using sums of global ratings.^2^ The clinical measures were used to assess symptoms of psychosis and mood dysregulation regardless of the diagnosis. Ratings on the BPRS and SANS/SAPS measures reflect symptom severity over the previous 30 days.

*Staff Training:* Staff training prior to independent administration of the diagnostic interview involved: reviewing the MINI and C-SSRS user guides and manuals, engaging in a group training and refresher meetings led by a PhD-level member of the team, completing practice ratings of interview videos, completing at least one mock interview with a fellow staff member, observing a trained rater administer at least two diagnostic interviews with study participants, and administering at least two diagnostic interviews with study participants while being observed by a trained rater. Staff training on symptom measures involved a similar process as the training for diagnostic interview administration (watching introductory videos provided by the scale authors that describe the interview tool, observing and co-rating, being observed by a trained staff member). The most recent inter-rater reliability check yielded an intraclass correlation of 0.92 across ten research staff rating the BPRS measure for a randomly selected sixteen participant interviews. Real-world functioning was measured using clinician ratings of participant engagement and performance in developmentally-appropriate social- and role-related activities at school, work, and home on the Global Functioning Social and Role Scales.^3,4^

*Behavioral Task: three-armed restless Bandit Task.* Prior to the task, participants completed 25 practice trials during which the reward probability of three choices was fixed at [0.7, 0.3, 0.2]. Two criteria needed to be met before proceeding with the task: 1. Participants needed to score above chance (at least 11 of 25 points), 2. Participants needed to switch choices at least twice. These criteria ensured that participant behavior was sufficiently flexible to explore options while also able to sustain reward. The practice training helped the participant to understand that the task is stochastic, such that even the best choice does not always provide a reward, and that they needed to do both exploring and exploiting to succeed in this task.

The walk (dynamic reward contingency of all three arms) was generated randomly with a few restrictions: (1) to ensure that no arm is constantly high payoff or low payoff, (2) to ensure the overall environmental richness was similar across walks, the average payoff probability across all three arms across the whole session was required to be 50%± 2%. (3) the reward probability was bound between 0.1 and 0.9 so the reward contingency was always stochastic and never deterministic (4) the initial reward probability of three arms was fixed at [0.9, 0.7, 0.3] for the first trial to ensure distinctiveness of choices at the beginning. The participants were instructed to collect as many points as possible. Participants were given minimal task instructions regarding the reward contingency to encourage naturalistic exploration behavior. They were informed only that reward contingencies would change slowly over time. No information was provided regarding starting reward probabilities, average payoff rates, or the speed or magnitude of changes. Thus, all aspects of task structure had to be inferred from trial-by-trial outcomes.

In each trial of the Bandit Task, participants were presented with three images of Gabor patches (i.e., sinusoidal luminance modulation within a Gaussian envelope, σ = 0.75°) on a mean gray background. Gabors differed in spatial frequency (0.75, 1.5 or 4 cycles/°) and orientation (45° apart), but were matched for contrast and color. The three Gabor images remained the same throughout the session but the location where each Gabor image appeared was randomized from trial to trial. The three images appeared at 4° eccentricity below-left, above-center, and below-right of the center fixation for 600 ms. Response was indicated by a left, center, or right key press on the task button-box during a 1600 ms response window (starting at the onset of the visual stimuli). The selected image was highlighted by a surrounding box around the selected stimulus for 400-800ms, randomly jittered by 100ms. Then the feedback to the selected choice was provided. The feedback was either +1 point (reward) or +0 points (reward omission) and was shown on the screen for a duration of 300-700ms, randomly jittered by 100ms. We used an empirically validated center fixation mark,^5^ which was also shown during the inter-trial interval with a duration of 500-1500ms, randomly jittered by 200ms. Stimuli were displayed using a BenQ PX9600 projector and viewed on a screen mounted at the back of the scanner through a mirror on the radiofrequency head coil. Projector luminance was linearized using a photometer. A four-button fiber optic response pad (Current Designs Inc. <https://www.curdes.com/> Philadelphia, PA, USA) was used to record participant responses. We used a Snellen eye chart^6^ to measure visual acuity and provided participants with MRI-safe corrective lenses when needed.

*Mixture of Exponential Distribution.* To examine whether participants’ choice behavior was characterized by a single strategy or multiple strategies with different time constants for switching, we analyzed the temporal structure of the choice sequences using a mixture model of exponential distributions. If choice behavior was underlied by a single time constant for switching, that is some fixed probability of moving away from the current option at any given trial, then we would expect to see the choice run durations (inter-switch intervals) be exponentially distributed:^7^

$$f(x) = \beta^{-1}e^{-\frac{x}{\beta}}$$

, where β is the scale parameter, which is related to the average inter-switch interval.

However, if there exist multiple time constants for switching in the choice sequence, the inter-switch intervals would be distributed as a mixture of multiple weighted exponential distributions:

$$f\left( x \right)=\sum_{i}^{n} \pi_{i}e^{-\frac{x}{\beta_{i}}}$$

, where β_i_ reflects the scale parameter (average inter-switch interval) for each component distribution, and π_i_ reflects the relative weight of each component. Therefore, the sum of all weights π_i_ is 1. Since inter-switch intervals are discrete, not continuous, we fit mixture models of 1-4 geometric distributions, which are the discrete equivalent of exponential distribution,^8^ via the expectation-maximization algorithm and calculated model log-likelihood for model comparison. Adding a second component distribution significantly improved model fit (**Figure S2A, B**, one component model log likelihood: control: -19233.8, EP: -21039.3, AIC: control: 38618, EP: 42214; mixture model of two components log likelihood: control: -17527.0, EP: -19400.5, AIC: control: 35354, EP: 39073). Although adding a third and fourth component further increased model likelihood (mixture model of three components log likelihood: control: -17440.8, EP: -19281, AIC: control: 35332, EP: 38970; mixture model of four components log likelihood: control: -17432.6, EP: -19274.5, AIC: control: 35465, EP: 39093), the second component granted the highest model performance improvement. Any additional components beyond the second one contributed only marginally to the log likelihood gains.^9^

In the mixture model fit to controls, the scaling parameter for the fast-switching regime, which is related to the switching time constant, is 1.8 trials, and the relative weight of this component distribution is 0.80; the time constant for the slow switching regime is 8.7 trials, and the relative weight of this component is 0.20. In the mixture model fit to EP, the time constant for the fast-switching regime is 1.7 trials, and the relative weight of this component distribution is 0.85; the time constant of the slow-switching regime is 7.8 trials and the relative weight of this component is 0.15.

*Hidden Markov Model (HMM):* Our HMM model consisted of two types of hidden strategy states, as defined by their unique emission probability of choices as well as transition probability between states. There is one explore state and three exploit states (“exploiting choice 1 state”, “exploiting choice 2 state”, “exploiting choice 3 state”). The exploit states only emit the choice that’s being exploited, e.g.: in exploit choice 1 state, the probability of choosing choice 1 is one, while the probability of choosing choice 2 and 3 is zero. The emissions model for the explore state was uniform across three options because this is the maximum entropy distribution for categorical variables. This does not require, imply, impose, or exclude that choices in explore states are random or structured.^10,11^ The latent states are assumed to be Markovian, meaning that states only depend on the most recent state. The transition matrix describes the one-step transition probabilities between each of the four states. The parameters were tied across the three exploit states, meaning that each exploit state had the same probability of transition into explore state and same probability of staying in exploit state. Transition probabilities from the explore state into one of the three exploit states were also tied. Our model also assumes that to transition from one exploit state to another exploit state, one must pass through the explore state, even just for one trial. Due to the independently changing reward contingency for each choice, when switching from exploiting one option to exploiting another, that very first trial should be exploratory as the reward contingency of the new option was unknown. Similarly, we initialized the first trial of the session in the explore state, as there was no prior knowledge about the reward contingency. The final transition matrix thus has two unique parameters - probability of transitioning from explore state to exploit state, and probability of transitioning from exploit state to explore state.

*Hidden Markov Model State Dynamics.* Further analysis of the transition matrix of the HMM can reveal the dynamics of choice behaviors. Previously, we have utilized analytical tools inspired from approaches used to understand physical mechanics and chemical kinetics to directly characterize the energetic landscape of behavior.^12^ We calculated the stationary distribution of the fitted HMMs, which is the equilibrium probability distribution over explore/exploit states. The stationary distribution is the relative frequency of being in an explore state and an exploit state that we would observe if the model’s dynamics were run for an infinite period of time. Since the Boltzmann distribution gives the probability that a system will be in a certain state as a function of that state's energy, the stationary distribution of the explore/exploit state allows us to derive the relative energy (depth) associated with each state.

To fully understand the energetic dynamics of the behavior, we also need to understand the activation energy required to transition between states, which is the height of the energetic barrier between two states. The Arrhenius equation relates the rate of transitions away from a state to the activation energy required to escape that state. This equation allows us to calculate the activation energy. Detailed equations can be found in the methods of our previous study.^11–13^

*Bayesian learner model (Kalman filter) with perseveration.* The Bayesian learner model assumes that one learns by consistently updating beliefs about the true underlying reward structure of the task - i.e. estimated reward distribution for each choice. On trial t, the model uses the previous feedback $r_{t-1}$ of selected choice $c_{t-1}$ to update the estimated value (EV) as well as the estimated variance $\sigma_{t}$, which reflects the uncertainty/confidence of the value estimates.

Since the restless bandit reward contingency is stochastic (average reward rate across all three arms is 0.5 ± 0.02) and dynamic (change frequency is 0.1, change magnitude is 0.1), we modeled a fixed observation noise parameter $\hat{\sigma}_{O}^{2}$ = 0.25 that represents the stochasticity in observed reward, and a fixed diffusion noise parameter $\hat{\sigma}_{D}^{2}$ = 0.1 that represents the task-specific reward diffusion process to distinguish from random decision noise. Both observation noise and diffusion noise were modeled as a Gaussian distribution with zero mean.

To allow for the learning of a changing reward structure, the model applied a decay parameter λ = 0.95 with decay center θ = 0.5 to “forget” learned expected value of each choice *c* on each trial *t*, according to: ${EV}_{c, t+1}=\lambda{EV}_{c,t}+(1-\lambda)\theta$. Similarly, the learned variance estimates also decay according to: $\hat{\sigma}_{c,t+1}^{2}= \lambda^{2}\hat{\sigma}_{c,t}^{2}+ \hat{\sigma}_{D}^{2}$. The intuition is that if a certain choice has not been selected over many trials, the learned value estimate of that choice will slowly decay toward random chance level (EV = 0.5), and the confidence in that learned value estimate also decreases, increasing the uncertainty. This is because the structure of the restless bandit task, where the payoff of each choice changes independently and randomly over time.

Participants update their value estimates of the chosen choice according to Bayes’ theorem. On the start of each trial, the prior belief about the reward distribution, which is normally distributed with mean reward rate $\hat{\mu}_{c,t}^{pre}$ and variance $\hat{\sigma}_{c,t}^{2 pre}$, is updated using prediction error $\delta_{c,t}=r_{t}-\hat{\mu}_{c,t}^{pre}$ by Kalman gain *k*_t_: $\hat{\mu}_{c,t}^{post}= \hat{\mu}_{c,t}^{pre}+ k_{t}\delta_{t}$ and $\hat{\sigma}_{c,t}^{2 post}=(1-k_{t})\hat{\sigma}_{c,t}^{2 pre}$.

The Kalman gain is similar to a learning rate in the reinforcement learning (RL) model in that it determines how much of the prediction error δ_t_ is being updated for value estimates. However, the Kalman gain depends on the estimated variance of the prior reward distribution and a fixed observation noise: $k_{t}= \hat{\sigma}_{c,t}^{2 pre}/(\hat{\sigma}_{c,t}^{2 pre}+\hat{\sigma}_{O}^{2}$ ).

The decay parameter (λ), which governs the rate at which unchosen options’ values and uncertainties revert toward baseline, was initially allowed to vary to capture potential individual differences in memory or uncertainty accumulation. Allowing λ to vary freely did not improve model evidence and did not reveal group differences in decay rate. Accordingly, λ was fixed in the final model to the empirical mean estimated from the free-decay model (λ = 0.95), reducing model complexity while retaining sensitivity to task dynamics.

The higher the estimated variance of reward distribution for choice *c* at trial *t*, the higher the Kalman gain. This means that the learning rate varies from trial to trial and is dependent on the uncertainty/confidence of the value estimate. When uncertainty is high (large estimated variance of reward distribution), the Kalman gain is high so that the immediate past feedback is weighted more and the reward history is weighted less, which promotes more exploration. When uncertainty is low, the Kalman gain is low so that the learned reward value from past trials is weighted more, which promotes more exploitation of a high value choice.

The model also included value-independent perseveration, which is the tendency to repeat the last choice regardless of the previous outcome, weighted by a perseveration weight ω during choice selection. With estimated expected value (${EV}_{c,t}=\hat{\mu}_{c,t}^{pre}$), estimated variance ($\hat{\sigma}_{c,t}^{2 pre}$) and perseveration, the choice selection was performed based on a SoftMax probability distribution:

$$p(c_{t+1})=\frac{e^{\beta({EV}_{c,t}+ \varphi\hat{\sigma}_{c,t}^{2 pre}+ I_{c_{t}=c_{t-1}}\omega)}}{\sum e^{\beta({EV}_{c,t}+ \varphi\hat{\sigma}_{c,t}^{2 pre}+ I_{c_{t}=c_{t-1}}\omega)}}$$

There are thus three free parameters in this model: uncertainty weight φ, perseveration weight ω, and inverse temperature β. φ denotes the uncertainty weight, which represents the degree to which estimated variance (uncertainty) influences choice selection. $I_{c_{t}=c_{t-1}}$is an indicator that equals 1 when current choice and previous choice are the same and ω is the perseveration weight, which reflects the degree to which the tendency to repeat the previous choice influences choice selection. Finally, inverse temperature β determines the level of random decision noise.

*Bayesian learner model (Kalman filter) with choice kernel.* We implemented an extended Bayesian learner model that incorporated a **choice kernel** that allows for choice repetition effects beyond trial t-1, analogous to *the Reinforcement learning + choice kernel (RLCK) model* below.

The learning component of this model was identical to the Bayesian learner (Kalman filter) model described above. In addition to value and uncertainty learning, the model included a **choice kernel (CK)** that captured value-independent choice repetition. The choice kernel tracked recent choice history for each option and was updated on each trial according to:

$${CK}_{t+1}^{k}={CK}_{t}^{k}+\alpha_{c}(I_{t}^{k}-{CK}_{t}^{k})$$

, where $I_{t}^{k}$ is an indicator variable equal to 1 if choice *k* was selected on trial *t*, and 0 otherwise. The $\alpha_{c}$ governs the rate at which recent choices influence future decisions. The choice selection was performed based on a SoftMax probability distribution:

$$p(c_{t+1})=\frac{e^{\beta({EV}_{c,t}+ \varphi\hat{\sigma}_{c,t}^{2 pre}+{CK}_{t+1}^{k})}}{\sum e^{\beta({EV}_{c,t}+ \varphi\hat{\sigma}_{c,t}^{2 pre}+ {CK}_{t+1}^{k})}}$$

*Reinforcement learning (RL) models.*

*Reinforcement learning model with first-order perseveration (RL+perseveration)*: To enable a more direct comparison with the Bayesian learner model, we implemented a reinforcement learning model that incorporates a **first-order perseveration term**. The model assumes that one learns by consistently updating an estimated value for each option at each trial t ($Q_{t}^{k}$).

In each trial, $r_{t}-Q_{t}^{k}$ captures the reward prediction error (RPE), which is the difference between expected value and the actual outcome. The parameter $\alpha$ is the learning rate, which determines the rate of updating RPE.

$$Q_{t+1}^{k}=Q_{t}^{k}+\alpha(r_{t}-Q_{t}^{k})$$

Choice selection was guided by a Softmax function that included a perseveration bias ω favoring repetition of the immediately preceding choice:

$$p\left( c_{t+1}=k \right)= \frac{e^{\beta{(Q}_{t}^{k}+I_{c_{t}=c_{t-1}}\omega)}}{\sum_{j} {(e}^{\beta{(Q}_{t}^{j}+I_{c_{t}=c_{t-1}}\omega)})}$$

$I_{c_{t}=c_{t-1}}$is an indicator that equals 1 when current choice and previous choice are the same and ω is the perseveration weight, which reflects the degree to which the tendency to repeat the previous choice influences choice selection. This formulation parallels the perseveration term in the Bayesian learner model, allowing for better comparison across models.

*Reinforcement learning + choice kernel (RLCK) model*: this model is a basic delta-rule reinforcement learning model with an additional choice kernel updating rule.

The choice kernel CK captures the value-independent tendency to repeat a biased choice. The choice kernel updating rule is similar to the value-updating rule:

$${CK}_{t+1}^{k}={CK}_{t}^{k}+\alpha_{c}(a_{t}^{k}-{CK}_{t}^{k})$$

Both value and choice kernel term were combined by a weight $\tau$ and used to guide decision making. The action selection was performed based on a Softmax probability distribution,

$$p\left( a_{t+1}=k \right)= \frac{e^{\beta{(\tau Q}_{t}^{k}+(1-\tau){CK}_{t}^{k})}}{\sum_{j} {(e}^{\beta{(\tau Q}_{t}^{j}+(1-\tau){CK}_{t}^{j})})}$$

where the inverse temperature $\beta$ determines how deterministic or stochastic the choice selection is based on the estimated values and choice kernel. The smaller the inverse temperature beta, the higher the decision noise; the larger the inverse temperature, the more reliance choice selection on estimated value and choice kernel.

*Asymmetric learning reinforcement learning (RL) + choice kernel model*. This model is an extension of the above model and incorporates an additional asymmetric learning scalar parameter that scales the learning rate on the trials where there is no reward, allowing for different learning rate for positive and negative prediction error. The choice selection is also based on a Softmax probability distribution where the inverse temperature determines the decision noise in the system.

$Q_{t+1}^{k}=\{Q_{t}^{k}+\alpha\left( r_{t}-Q_{t}^{k} \right),r_{t}=1 Q_{t}^{k}+\gamma\times\alpha\left( r_{t}-Q_{t}^{k} \right),r_{t}=0$

*Dual state reinforcement learning (RL) model.* Since learning rate might be different during explore state and exploit state, we also fitted a dual-state RL model. This model is largely the same as the RLCK model, except this model assumes two learning rates for HMM-inferred explore and exploit state. $\gamma$ scales learning rate during exploit state.

$$\{Q_{t+1}^{k}= Q_{t}^{k}+\alpha\left( r_{t}-Q_{t}^{k} \right), state=explore Q_{t+1}^{k}= Q_{t}^{k}+\gamma*\alpha\left( r_{t}-Q_{t}^{k} \right), state=exploit$$

AIC and BIC values of each model for each group were calculated for model comparison to determine the best model with the highest relative likelihood. For the control group, Bayesian learner model + perseveration: AIC = 43124.82, BIC = 44254.78; Bayesian learner model + choice kernel: AIC = 42526.11, BIC = 43656.06; RL + perseveration: AIC = 43526.63, BIC = 44656.58; RL + choice kernel: AIC = 45746.44, BIC = 46205.04; asymmetrical RL + choice kernel: AIC = 42967.11, BIC = 44850.36; dual-state RLCK model: AIC = 45487.09, BIC = 46060.35. For the early psychosis group: Bayesian learner model + perseveration: AIC = 49600.68, BIC = 50789.04; Bayesian learner model + choice kernel: AIC = 50063.58, BIC = 51251.94; RL + perseveration: AIC = 49697.38, BIC = 50885.75; RL + choice kernel: AIC = 51684.88, BIC = 52181.36; asymmetrical RL + choice kernel: AIC = 49320.36, BIC = 51300.97; dual-state RLCK model: AIC = 51429.96, BIC = 52050.57.

We selected the *Bayesian learner + perseveration model* as the primary model for inference. This choice was motivated by (i) parsimony and interpretability, and (ii) its superior performance in the early psychosis group under BIC, which imposes a stronger penalty for additional parameters and therefore favors models that generalize rather than overfit. Although AIC slightly favored the asymmetrical RL + choice-kernel model in early psychosis, this advantage was not retained under BIC. We therefore treated BIC as the primary criterion for model selection, while reporting AIC results to document sensitivity to the choice of information criterion. For completeness, we note that controls were best fit by the Bayesian learner + choice-kernel model; however, we prioritized a common, parsimonious model for downstream parameter-based analyses focused on early psychosis.

*Principal Component Analysis (PCA).* We performed a Principal Component Analysis (PCA) to reduce the dimensionality of all behavioral data and identify key components contributing to variance in the dataset (**Figure S6**). Five behavioral features were included for analysis: two key computational parameters extracted from the bandit task (uncertainty weight and inverse temperature) and three key cognitive measures from TestMyBrain (processing speed accuracy, verbal memory accuracy, and executive function accuracy). PCA was then applied to the standardized data to ensure all variables contributed equally, irrespective of their original scales. The explained variance ratio for each principal component was calculated to assess the proportion of variance captured. Additionally, the PCA loadings (feature contributions to each principal component) were extracted to interpret the relationship between the original features and the principal components. To assess the significance of feature loadings on each PC, we conducted a permutation test by shuffling the data 1000 times, recalculating the PCA for each permutation, and comparing the observed loadings to null distributions.

**Supplementary Results.**

Results across session 1 and session 2 are qualitatively similar, and are presented for side-by-side comparison in **Figures S1-2, 4**.

Apart from similar reward acquisition performance, additional evidence of equal task engagement was that we found no group differences in the number of trials completed in either session (*F*(1,141)=1.68, *p*=.197), nor in stimulus position bias (repeated key press) (*p*>.302 at session 1, *p*>.263 at session 2) and only a non-significant trend toward a group difference in response time (time elapsed between stimulus onset and key press for response), consistent with moderate slowing on timed tests among people with psychosis ^14^ (*F*(1,141)=3.42, *p*=.067). The similar response time across groups suggests that the less “optimal” choice selection in participants with EP was not a speed accuracy tradeoff ^15^.

The statistical analysis results presented in the main text account for session time points. Notably, even though the reward contingency (“walk”) was generated and assigned to participants randomly, chance level of reward under each walk differed significantly between groups at session 1 (*F*(1,141)=6.04, *p*=.015) such that random chance of getting a reward was slightly higher for participants with EP (mean = 50.07%, SD=0.75) than control participants (mean = 49.78%, SD=0.65). This would make it slightly more difficult to perform above chance for participants with EP. However, all chance levels of obtaining reward were fixed within a range from 48-52%, inclusive, and chance level of reward was not significantly different between groups at session 2 (*F*(1,122)=0.03, *p*=.860). Thus, this unintentional group difference in walks at session 1, while statistically significant, is not a meaningful difference.

To examine whether participants’ choice behavior was characterized by a single strategy or multiple strategies with different time constants for switching, we analyzed the temporal structure of the choice sequences using a mixture model of exponential distributions. If choice behavior had a single underlying time constant for switching, or, in other words, some fixed probability of moving away from the current option at any given trial, then we would expect to see the inter-switch intervals (the number of consecutive repeated choices between switches) be exponentially distributed ^7^. However, if there exist multiple time constants for switching in the choice sequence, the inter-switch intervals would be distributed as a mixture of multiple weighted exponential distributions. A mixture of two components — a fast-switching regime (putatively exploration) and a slow-switching regime (putatively exploitation) — provided the most parsimonious explanation for the choice behaviors in both control and EP groups (**Figure 2A, 2B**), consistent with the performance of non-human animals (mice ^11,12^, and monkeys ^10,12^) on equivalent tasks.

We observed higher exploration rates across both sessions for EP compared to the control group. Specifically, EP had 36.7% ± 27.1% of trials labeled as exploration at session 1 and 32.0% ± 22.3% at session 2, whereas controls had 27.4% ± 18.3% of trials labeled as exploration at session 1 and 23.8% ± 16.6% at session 2 (**Figure S2A**).

The model-labeled strategy states were also associated with different win-stay lose-shift patterns, which capture the observed changing sensitivity to outcomes. We found a higher probability of win-stay during the exploit state than during the explore state (main effect of state, *F*(385)=137.73, *p*<.001) and higher probability of lose-shift during the explore state than during the exploit state (main effect of state, *F*(385)=2100.48, *p*<.001) in both groups, implicating adaptive outcome sensitivity in explore and exploit strategy state and supporting the face validity of HMM-state labels to capture meaningfully distinct latent strategies.

Further analysis of the transition matrix can reveal the dynamics of choice behaviors and provide an intuitive way to visualize the stability of strategy states. We derived the relative energy associated with the explore/exploit state from the stationary distribution of two strategy states and calculated the activation energy required to move from one state to another (See Methods) (**Figure S2F**). The stationary distribution of explore/exploit strategy states characterize the stability of each strategy state. The deeper the basin of a state, the more stable the state is. The shallower the state, the less stable it is. The explore and the exploit state each have some inertia, i.e.: some probability of staying in that state. The activation energy of each strategy state characterizes the energy required to transition out of that strategy and into the other strategy. The harder it is to transition out of a strategy state, the higher the height of the energetic barrier. In both control participants and participants with EP, the depth of the exploit state basin was deeper than the explore state, suggesting that exploitation is a more stable strategy state than exploration. Furthermore, in models fit to EP, exploitation was a less stable behavioral state in EP (stationary probability of exploration, EP: 60.0% ± 20.0% STD), compared to models fit to controls (stationary probability of exploration, control: 67.0% ± 16.6%, different from EP: *p*=.0024, 95% CI for the difference = [-11.4%, -2.5%]).

**Model comparison: A Bayesian learner model that adjusts learning rate based on uncertainty estimates explained EP participant choice behaviors better than RL models with fixed learning rates**

Our behavioral analyses suggested that excessive switching during exploitation was not explained by simple reward learning deficits. In particular, we did not observe group differences in the probability of switching during exploration or in the probability of transitioning into exploitation. This pattern raised the possibility that exploration behavior may be driven not only by reward learning but also by uncertainty associated with unchosen options. To examine this possibility, we turned to computational models that allow us to dissociate latent decision processes contributing to exploration.

We first implemented a Bayesian learner model that utilized Kalman Filters to fit participants' choices. Recent work suggests that the Bayesian learning framework is especially well-suited for dynamic tasks like ours, where volatility in reward contingencies challenges individuals to continuously update their beliefs and adapt their strategies ^16–19^. This framework explicitly models how individuals adjust their learning rates based on the level of uncertainty in the environment, which has been shown to be relevant in populations with psychosis ^20^. In this framework, the effective learning rate (Kalman gain) adapts dynamically according to the uncertainty associated with value estimates, placing greater weight on new observations when uncertainty is high and relying more on accumulated reward history when uncertainty is low. Because the restless bandit task involves stochastic and evolving reward contingencies, this framework allows us to examine how individuals update both expected value and confidence in those estimates over time. Choice selection in this model depends on both estimated value and estimated uncertainty through a SoftMax decision rule. To account for value-independent choice repetition, we included a first-order perseveration term.

For completeness, we also considered another prominent class of computational models of value-based decision making, that of reinforcement learning (RL). The RL framework describes a process of value updating using feedback ^21,22^. RL models are widely used in modeling Bandit task behaviors. However, the learning rates in RL models are often fixed or differ only between specific strategy states. Here, we included RL models as an important comparison because the RL framework provides a robust and well-established baseline to determine whether participants’ decisions can be explained primarily by value-driven learning mechanisms or if there is a unique contribution of uncertainty estimates that the Bayesian approach captures. We therefore evaluated several RL variants commonly used in the literature, including models with fixed learning rates, asymmetric learning rates for positive and negative prediction errors, and dual-state RL models with separate learning rates during exploration and exploitation states^11,23–25^. Because restless bandit behavior often exhibits value-independent choice persistence, we also implemented alternative formulations of choice repetition using either a first-order perseveration term or a choice kernel that captures longer-range choice history effects.

Model comparison was performed using Akaike Information Criterion (AIC) and Bayesian Information Criterion (BIC). Across models, the Bayesian learner model with first-order perseveration provided the best fit to Early Psychosis participant behavior under BIC in both groups (**Figure 3A**). This result suggests that allowing learning rates to adapt dynamically based on uncertainty provides a better account of choice behavior of Early Psychosis individuals in the present restless bandit task than RL models with fixed learning rates.

#### Sensitivity analysis: relaxing the uncertainty-weight constraint

To evaluate whether constraining the uncertainty weight parameter (φ) to be non-negative influenced model fit or group-level inferences, we fit an alternative Kalman filter model in which φ was unconstrained and allowed to take negative values. This specification permits the same uncertainty signal to either promote exploration (φ>0) or discourage sampling of uncertain options (φ<0), corresponding to uncertainty avoidance.

Allowing φ to vary freely did not improve model fit in the early psychosis (EP) group. Instead, the unconstrained model yielded worse negative log-likelihood relative to the constrained model, despite its additional flexibility. Group comparisons under this unconstrained specification showed an attenuated difference in φ between EP and controls (p = 0.13), indicating that relaxing the positivity constraint did not strengthen evidence for group differences. However, parameter recovery analyses indicated that when φ was allowed to take negative values, the recovery of all parameters deteriorated substantially, reflecting strong parameter trade-offs between uncertainty weighting, decision noise, and perseveration in this regime. Thus, relaxing the positivity constraint did not strengthen evidence for group differences and did not improve descriptive adequacy for EP behavior.

#### Parameter recovery under negative uncertainty-weight regimes

To assess whether uncertainty avoidance (negative uncertainty weighting) is practically identifiable in the present task, we conducted targeted parameter recovery analyses in which simulated datasets were generated using a Kalman filter model with uncertainty weight φ sampled uniformly from a negative range (φ ∈ [−10, −0.01]). All other parameters were sampled from the same ranges used in the primary recovery analyses, and simulated data were fit using the identical estimation pipeline (MLE with multi-start optimization).

Recovery was substantially reduced in this negative-φ regime. Specifically, recovery of φ was markedly weaker (corr = 0.64; RMSE = 3.24; bias = −1.04), and recovery of β (corr = 0.85; RMSE = 2.32; bias = 1.26) and ω (corr = 0.43; RMSE = 0.39; bias = 0.15) was also degraded relative to the constrained (φ ≥ 0) model. This pattern indicates increased parameter tradeoffs and reduced practical separability among uncertainty weighting, choice stochasticity, and perseveration when φ is allowed to take negative values.

Together with the poorer fit of the unconstrained model to early psychosis behavior, these findings suggest that modeling uncertainty avoidance as a negative uncertainty bonus is not well supported by the present task and is difficult to dissociate from other mechanisms in finite data. Accordingly, we retain the constrained formulation (φ ≥ 0) in the primary analyses and interpret φ as indexing the strength of an uncertainty bonus (directed exploration), with φ ≈ 0 reflecting minimal uncertainty-guided exploration.

**Uncertainty sensitivity and decision noise both contribute to changes in exploration-exploitation tradeoff**

To determine how changes in the Bayesian learner model parameters influence the level of HMM-inferred exploration, we ran computer simulations of 10,000 Bayesian learner agents defined by different combinations of two main free parameters: uncertainty weight (φ) and inverse temperature (β) doing a three-armed restless bandit task. We fit a Hidden Markov model (HMM) to the simulated choice sequence and inferred each choice as being in an explore or exploit state. Then we plotted the level of exploration produced by agents with varying uncertainty weight and inverse temperature parameters (**Figure 3B**). As expected, changes in both uncertainty weight and inverse temperature influenced the level of exploration in the restless bandit task. Because higher uncertainty weight reflects greater uncertainty sensitivity and lower inverse temperature reflects greater decision noise, we report our results using these descriptive labels throughout (uncertainty sensitivity for φ and decision noise for β). Exploration increased with both higher uncertainty sensitivity and with higher decision noise (**Figure 3B**).

**Uncertainty sensitivity and decision noise account for interindividual variance in cognitive processes independent of traditional cognitive measures**

In order to determine whether the computational parameters extracted from our task identify relevant cognitive processes beyond traditional measures, we created a vector out of computational parameters from the bandit task (uncertainty sensitivity and decision noise) and traditional cognitive scores (processing speed accuracy, verbal memory accuracy, and executive function accuracy) from TestMyBrain measures of processing speed (digit symbol matching test), executive function (matrix reasoning test), and verbal memory (verbal paired associates test) ^26^; we conducted a Principal Component Analysis (PCA) to examine the major axes that capture interindividual variability in cognitive processing. The purpose of the PCA was not to reduce dimensionality but to identify the unique contributions of the various cognitive features to interindividual variability. PCA was applied to these five features (after z-scoring) to identify latent components that capture shared variance. This yielded five interpretable principal components (PCs), with the first three PCs explaining the majority of the variance in cognitive processing (~ 76.5%) (**Figure S6A,** See Supplement). To examine what each PC represents, we analyzed feature loadings to determine the correlation of each computational and cognitive variable to the principal components (**Figure S6B**).

The first principal component (PC1) captured ~38% of the variance and loaded moderately on decision noise (loading = 0.41, *p* = .54) and all traditional cognitive measures, including processing speed accuracy (loading = 0.58, *p* = .21), verbal memory accuracy (loading = 0.46, *p* = .47), and executive function accuracy (loading = 0.54, *p* =.29). However, none of these loadings were statistically significant, suggesting that PC1 represents a general cognitive factor encompassing shared variance across multiple domains. The second principal component (PC2), which explained ~23% of the variance, was strongly dominated by uncertainty sensitivity (loading = 0.83, *p* = 0.009). Notably, no other feature contributed significantly to PC2. The third principal component (PC3), which explained ~16% of the variance, specifically captured variance related to decision noise (loading = 0.86, *p* = .03) and similarly, traditional cognitive measures contributed minimally to this PC and were not statistically significant. Our findings from PC2 and PC3 indicate that uncertainty sensitivity and decision noise each identify a distinct and independent microcognitive process that is orthogonal to traditional cognitive measures. The remaining variance accounted for by PC4 and PC5 was dominated by verbal memory accuracy (loading = 0.74, *p* = 0.051) and processing speed accuracy (loading = 0.77, *p* = .001) respectively, reflecting unique contributions from these cognitive domains. Together, PCA revealed that the computational parameters derived from the task, specifically uncertainty sensitivity (PC2) and decision noise (PC3), captured distinct and independent components of interindividual cognitive variance that are not represented by traditional measures of cognitive ability.

**Figure S7** shows the test-retest reliability of the computational parameters of the task, which was estimated with a two-way consistency intraclass correlation model. Figure S7E shows data after Winsorizing four uncertainty sensitivity parameter outliers across both sessions (to the value of the 99th percentile in the distribution). Reported results used winsorized values for the uncertainty sensitivity parameter. Notably, while there were individual differences in model fit, results were robust and not driven by outliers.

We examined whether individual characteristics such as gender identity explained task strategy differences across participant groups. There was a significant group-by-gender interaction effect for probability of exploration (**Figure S8**, *F*(2,139)=3.96, *p*=.021) and uncertainty sensitivity (*F*(2,139)=3.78, *p*=.025). Pairwise comparisons revealed that cis-men participants with EP had the highest probability of exploration and uncertainty sensitivity compared to: cis-men control participants (*p*<.001, *p*<.001), cis-women participants with EP (*p*<.001, *p*<.001) and nonbinary participants with EP (*p*=.002; *p*<.001, respectively). Thus, participants with EP identifying as cis-men had the highest probability of exploration and uncertainty sensitivity, suggesting that these participants are most likely to engage in the computational failures that are detected by this task.

Given a group-by-gender interaction effect in predicting task strategy, we further investigated individual differences. Interestingly, there was a significant interaction effect between number of lifetime psychiatric hospitalizations and gender for predicting uncertainty sensitivity (*F*(4,135)=2.79, *p*=.029) such that cis-men participants with EP who had multiple lifetime psychiatric hospitalizations had the highest uncertainty sensitivity compared to: cis-women participants with EP who had multiple hospitalizations (*p*<.001), nonbinary participants with EP who had multiple hospitalizations (*p*<.001), cis-men participants with EP who had 0 or 1 lifetime psychiatric hospitalization (*p*=.047), and cis-men control participants (*p*<.001). Similarly, there was a gender-by-lifetime psychiatric hospitalization interaction that trended toward significance in predicting probability of exploration (*F*(4,135)=2.25, *p*=.067). This suggests that individual differences in gender identity and lifetime psychiatric hospitalizations, which may represent a proxy for life stressors or volatility in the participant’s environment, may contribute to task strategy.

The gender effect on exploration was not explained by current symptom severity (BPRS total; *F*(2,71)=2.38, *p*=.100), psychotic illness duration (*F*(2,72)=0.33, *p*=.723), or antipsychotic medication status (𝜒^2^(2,75)=5.03, *p*=.081) when comparing cis-men with EP to other gender identities among participants with EP. Further, there was no evidence of a general pattern of over-exploration among cis-men, based on a non-significant effect of gender on exploration among controls only (*F*(2,66)=1.08, *p*=.344). However, there was a significant gender difference in diagnostic group composition such that cis-women were overrepresented in the affective psychosis group compared to the non-affective psychosis group among EP (𝜒^2^(2,75)=8.52, *p*=.014). We therefore compared affective vs. non-affective psychosis vs. control groups and found a gender-by-diagnostic group interaction effect trending towards significance for probability of exploration (**Figure S8**, *F*(4,136)=2.09, *p*=.085) such that cis-men with non-affective psychosis (*p*=.007) and cis-men with affective psychosis (*p*=.006) were more exploratory than cis-men in the control group. Within the non-affective psychosis group, cis-men were more exploratory than cis-women (*p*=.013) and nonbinary participants (*p*=.023). Within the affective psychosis group, cis-men were more exploratory than cis-women (*p*=.002) and nonbinary participants (*p*=.092). Similarly, a gender-by-diagnostic group interaction for uncertainty sensitivity trended towards significance (*F*(4,136)=2.23, *p*=.069). Notably, antipsychotic medication status was equally distributed across affective and non-affective groups (𝜒^2^(1,74)=0.13, *p*=.719). Finally, there was no significant effect of age, or age-by-group interaction, on computational task parameters (*p*>.084). Together, these results suggest a link between individual differences in task strategy, gender identity, and lifetime psychiatric hospitalizations that was not driven by clinical characteristics (symptom severity, duration of psychotic illness, diagnostic subgroups, antipsychotic medication).

**Figure S9** shows details of the cluster analysis metrics, suggesting that three separate clusters best represent the data. Table S2 provides the ANOVA results showing how the three clusters differed on strategy. To limit Type I error, these analyses were conducted on the cluster variable rather than on group by cluster.

**Supplementary Discussion.**

Examining the influence of gender groups and clinical characteristics adds another layer of complexity. We found that over-exploration was driven by participants with EP who identified as cis-men, regardless of whether in the affective or non-affective psychosis group. We also found that overweighting of uncertainty, which partially explained group differences in exploration, was driven by participants with EP with multiple hospitalizations as compared to those with one or none. Thus, individual differences in gender identity and lifetime psychiatric hospitalizations, which may represent a proxy for life stressors or volatility in the participant’s environment, may contribute to task strategy. This is consistent with previous work suggesting that there are subtypes of people with psychosis who have different patterns of exploration ^27^. Our participants were relatively young, with relatively recent psychosis onset (median duration of psychosis was three and a half years). Previous research suggests that adolescence is characterized by enhanced novelty seeking and response shifting especially after negative feedback^28^. Further, directed, information-seeking, exploration increases during typical development in adolescence ^29^. It is possible that participants with EP in our sample, with an average age of 25, have not yet matured past this stage of typical development. Alternatively, it is possible that their development of more adaptive strategies was interrupted by the illness onset during adolescence/early adulthood. We did not observe an effect of age, or an age-by-group interaction, on computational task parameters, suggesting the latter interpretation. This implies that participants with EP have an altered developmental trajectory of value-based decision making strategy that warrants clinical attention. Future longitudinal work is needed to determine how task strategy changes over time and with varying clinical trajectories and interventions during this early, neuroplastic phase of illness.

#### Supplementary Tables and Figures.

#### Table S1. Participant Characteristics

| **Mean (SD) or %** | **Control**  **n=68** | **Early Psychosis (EP)**  **n=75** | **Statistic** |
| --- | --- | --- | --- |
| **Age** | 26.5 (6.8) | 25.0 (5.1) | *t*(124.57)=1.47, *p*=.145 |
| **Gender: % cis-woman, cis-man, other** | 54.4, 39.7, 5.9 | 38.7, 29.3, 32.0 | *χ*^2^(2,143)=15.46, *p*<.001 |
| **Racial Identity: % White, Black, Indigenous American, Asian, Multiracial** | 72.1, 2.9,  0, 16.2, 8.8 | 70.7, 9.3,  2.7, 6.7, 10.7 | *χ*^2^(4,143)=7.15, *p*=.128 |
| **Parent Education, years** | 15.5 (3.4) | 14.6 (3.0) | *t*(141)=1.73, *p*=.086 |
| **Estimated IQ** | 101.7 (8.3) | 99.3 (9.8) | *t*(137)=1.50, *p*=.136 |
| **Symptom Severity, BPRS subscales** |  |  |  |
| **Positive symptoms** | 5.1 (0.3) | 9.9 (5.0) | *t*(74.37)=-8.32, *p*<.0001 |
| **Negative symptoms** | 3.6 (1.0) | 5.5 (3.2) | *t*(89.19)=-4.96, *p*<.0001 |
| **Disorganized symptoms** | 4.9 (1.2) | 6.9 (2.0) | *t*(123.9)=-7.34, *p*<.0001 |
| **Depression symptoms** | 4.2 (1.5) | 8.6 (3.7) | *t*(98.88)=-9.66, *p*<.0001 |
| **Mania symptoms** | 3.2 (0.6) | 4.0 (1.9) | *t*(90.82)=-3.39, *p*=.001 |
| **Global Functioning, role** | 8.5 (0.9) | 6.6 (1.4) | *t*(123.18)=10.05, *p*<.001 |
| **Global Functioning, social** | 8.5 (1.0) | 7.1 (1.6) | *t*(125.85)=6.24, *p*<.001 |
| **Repeat session completed** | 92.6% | 81.3% | N/A |
| **Time between sessions (days)** | 22.8 (23.6) | 23.0 (30.0) | *t*(114.01)=-0.06, *p*=.95 |
| **Diagnoses** | N/A | 30.7% schizophrenia,  14.7% schizoaffective,  28.0% bipolar disorder with psychotic features,  10.7% major depression with psychotic features,  16.0% other psychosis | N/A |
| **Duration of psychotic illness, years** | N/A | 5.0 (4.5) | N/A |
| **Antipsychotic medication load** | N/A | 561.4 (1185.6) | N/A |

* Note: Gender “other” includes all non-cis gender identities such as trans and non-binary; BPRS = Brief Psychiatric Rating Scale, minimum scores per symptom subscale: positive=5, negative=3, disorganized=4, depression=3, mania=3; Global Functioning ranges from 1(extreme dysfunction) - 10(superior functioning) with 8=good functioning and 7=mild impairment; Diagnoses “other psychosis” category includes other psychosis spectrum disorders such as schizophreniform, psychosis not otherwise specified, and delusional disorder; For analyses, the non-affective psychosis group included diagnoses of schizophrenia, schizoaffective, and other psychosis, whereas the affective psychosis group included bipolar disorder and major depression with psychotic features. Antipsychotic medication load was estimated by calculating chlorpromazine equivalents based on the Defined Daily Dose method ^30^; Missing data due to data loss: 1 control participant and 3 participants with EP on IQ, 1 participant with EP on symptom ratings (BPRS) due to interruption of video function during interview that prevented ratings based on observation, 1 participant with EP on global functioning ratings.

**Table S2. Parameter recovery for Bayesian Learner model**

| **Parameter** | **Model specification** | **Recovery r** | **Bias (mean)** | **RMSE** |
| --- | --- | --- | --- | --- |
| β | Winning model (φ > 0) | 0.964 | 0.187 | 0.802 |
| φ | Winning model (φ > 0) | 0.921 | −0.052 | 1.209 |
| ω | Winning model (φ > 0) | 0.925 | −0.001 | 0.113 |
| β | Alternative model (φ unconstrained) | 0.924 | 0.419 | 1.268 |
| φ | Alternative model (φ unconstrained) | 0.916 | 0.070 | 1.278 |
| ω | Alternative model (φ unconstrained) | 0.810 | 0.017 | 0.190 |
| β | Alternative model (φ < 0) | 0.846 | 1.263 | 2.315 |
| φ | Alternative model (φ < 0) | 0.637 | −1.041 | 3.240 |
| ω | Alternative model (φ < 0) | 0.431 | 0.150 | 0.388 |

**Table S3. Two-Way ANOVA results and FDR-Corrected Pairwise Comparisons** (related to Figure 4D-F)

**ANOVA main effect of subtype**

| **Variable** |  | **F-Statistic** | **p-Value** |
| --- | --- | --- | --- |
| Level of exploration |  | 12.093 | <.001 |
| Transition to explore |  | 12.094 | <.001 |
| Transition to exploit |  | 4.617 | .011 |

### **FDR-Corrected Pairwise Comparisons**

#### Level of exploration (p(explore))

| *Group A* | *Group B* | *t* | *p-uncorrected* | *p-corrected* |
| --- | --- | --- | --- | --- |
| **1** | **2** | **-3.270** | **.002** | **.003** |
| **1** | **3** | **-3.930** | **<.001** | **<.001** |
| 2 | 3 | -.713 | .478 | .478 |

#### Transition to exploration

| *Group A* | *Group B* | *t* | *p-uncorrected* | *p-corrected* |
| --- | --- | --- | --- | --- |
| **1** | **2** | **6.845** | **<.001** | **<.001** |
| **1** | **3** | **4.660** | **<.001** | **<.001** |
| 2 | 3 | -.010 | .99 | .99 |

#### Transition to exploitation

| *Group A* | *Group B* | *t* | *p-uncorrected* | *p-corrected* |
| --- | --- | --- | --- | --- |
| 1 | 2 | .793 | .431 | .431 |
| **1** | **3** | **-2.16** | **.032** | **.048** |
| **2** | **3** | **-2.130** | **.023** | **.048** |

**Table S4. Spearman correlations with FDR-correction for multiple comparisons.** (related to Figure 5)

|  | *1* | *2* | *3* | *4* | *5* | *6* | *7* | | *8* | | *9* |
| --- | --- | --- | --- | --- | --- | --- | --- | --- | --- | --- | --- |
| *1. BPRS positive* | -- |  |  |  |  |  | |  | |  |  |
| *2. BPRS negative* | .23 | -- |  |  |  |  | |  | |  |  |
| *3. BPRS disorganized* | **.56***** | .27 | -- |  |  |  | |  | |  |  |
| *4. BPRS mania* | .30 | -.16 | .08 | -- |  |  | |  | |  |  |
| *5. BPRS depression* | **.42**** | .09 | .31 | .09 | -- |  | |  | |  |  |
| *6. Role functioning* | -.21 | -.25 | -.32 | -.09 | -.08 | -- | |  | |  |  |
| *7. Transition to explore* | .01 | .34 | .03 | -.17 | -.18 | **-.40*** | | -- | |  |  |
| *8. Uncertainty sensitivity* | -.16 | .14 | -.17 | -.07 | -.12 | .03 | | .09 | | -- |  |
| *9. Decision noise* | -.04 | **-.37*** | -.10 | .10 | .17 | .36 | | **-.77***** | | .21 | -- |

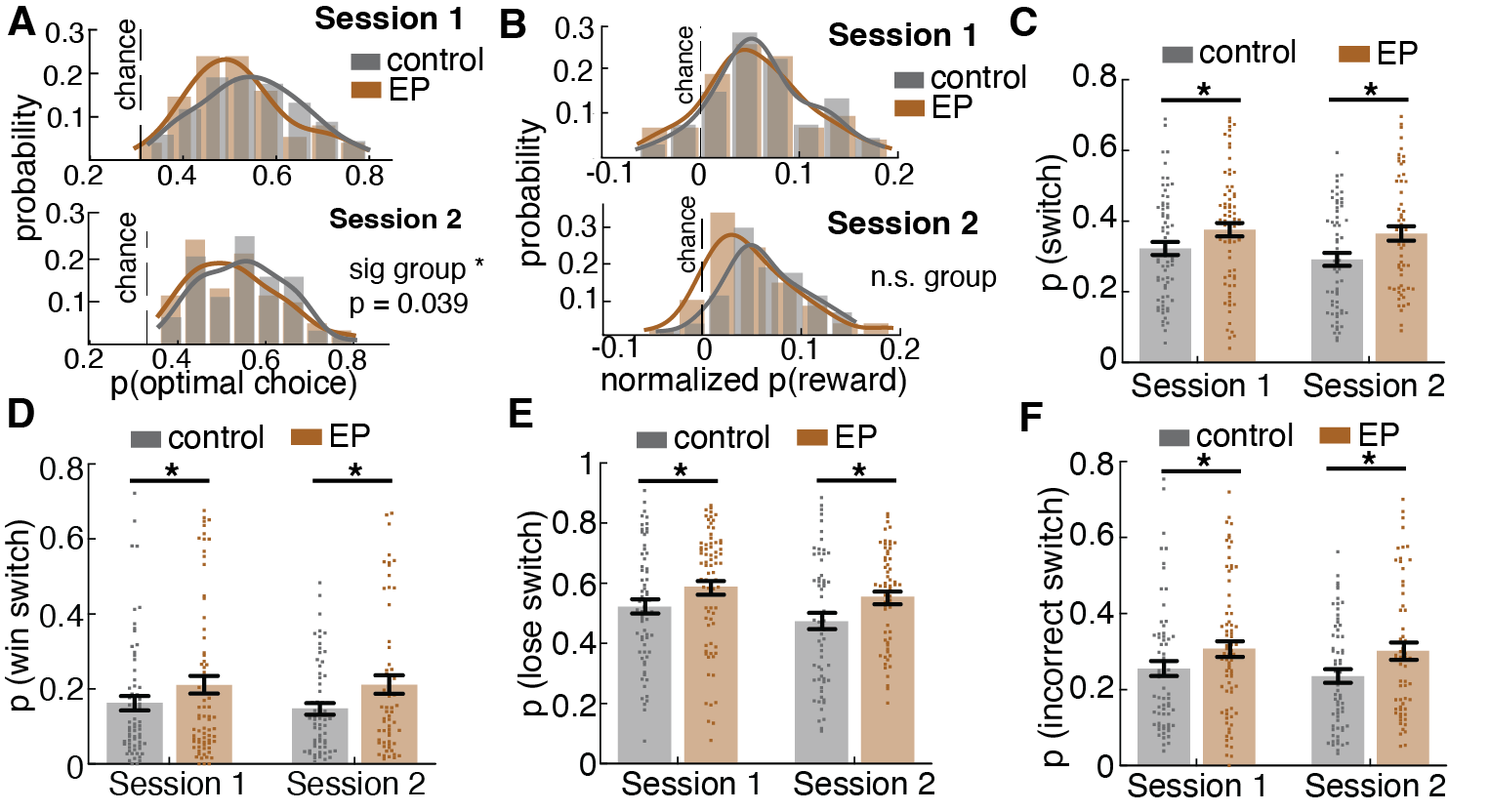

**Figure S1. Task performance and switching behaviors in the restless bandit task across Session 1 and Session 2. Related to Figure 1. A**) Average probability of choosing the optimal choice (highest payoff choice), **B**) Average reward acquisition normalized by chance level, **C**) Average probability of switching to a different choice, **D**) Average probability of switch after a rewarded trial (win switch), **E**) Average probability of switch after a reward omission trial (lose switch), **F**) Average probability of incorrect switch (switching away from an optimal choice).

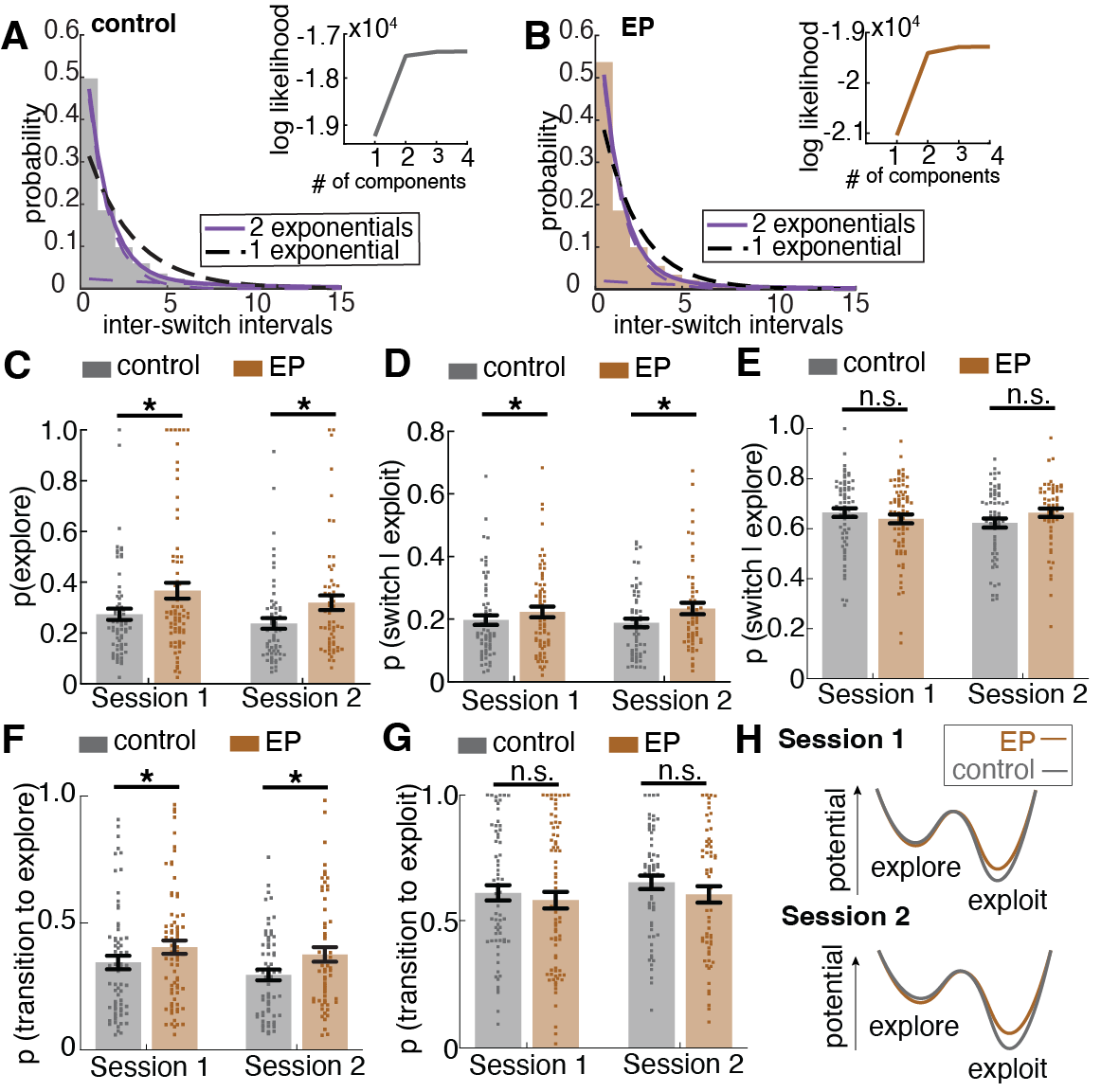

**Figure S2. Hidden Markov Model (HMM)-inferred exploration and explore/exploit transition dynamics across Session 1 and Session 2. Related to Figure 2. A, B**) The distribution of inter-switch intervals and mixture model fit for control participants (A) and participants with EP (B). A single probability of switching would produce exponentially distributed inter-switch intervals. Black dotted line - the maximum likelihood fit for a single discrete exponential distribution. Purple solid line - a mixture of two exponential distributions, with each component distribution in dotted purple line. The two components reflect one fast-switching regime and one persistent regime. The inset is the log likelihood of mixtures of 1-4 exponential distributions. A clear elbow effect can be observed at 2 components. **C**) Probability of exploration inferred from HMM, **D**) Probability of switching during exploit strategy state, **E**) Probability of switching during explore strategy state, **D**) Probability of transitioning to explore strategy state from exploit strategy state, **G**) Probability of transitioning to exploit strategy state from explore strategy state, **H**) Dynamic landscape of fitted HMMs for controls and EPs for session 1 (top) and session 2 (bottom). The model fit to EP had shallower and less stable exploit state, making it easier to transition out of exploiting a high value choice and start exploration too soon.

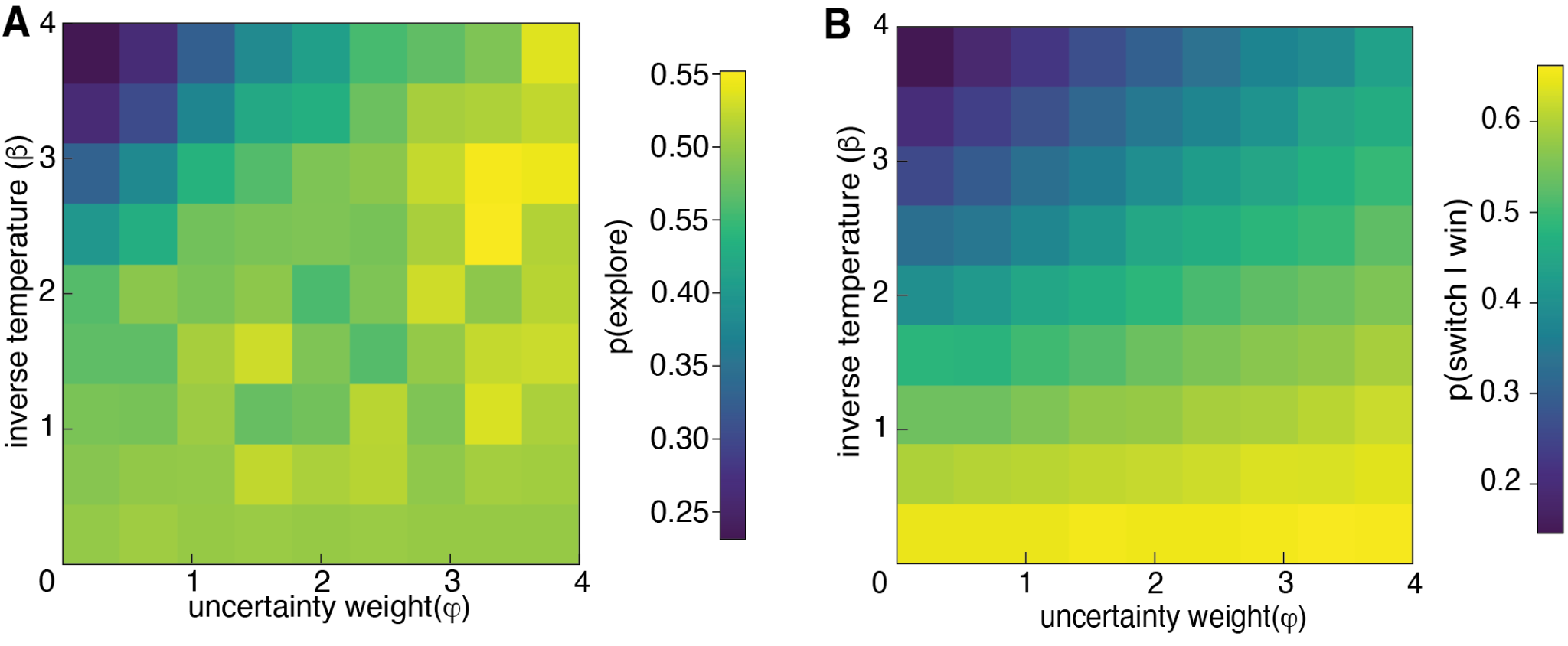

**Figure S3. A Bayesian Learner model simulated exploration and win-switching behavior in the restless bandit task. A)** Simulation of 10,000 Bayesian learner agents with different random combinations of inverse temperatures (β) and uncertainty weight (φ), performing the Translation Bandit task and HMMs were fitted to simulated choice sequences to infer the level of exploration. Heatmap of all pairwise combinations of inverse temperature and uncertainty weight. The color of the heatmap represents the level of HMM-inferred exploration. High uncertainty weight or low inverse temperature (high decision noise) leads to a high level of exploration. **B)** Heatmap illustrating the probability of switching after a rewarded outcome (p(switch|win)) across the same range of β and φ values. Elevated win-switching emerges from the joint influence of higher uncertainty sensitivity (φ) and increased decision noise(β^-1^) (n_sim = 10,000).

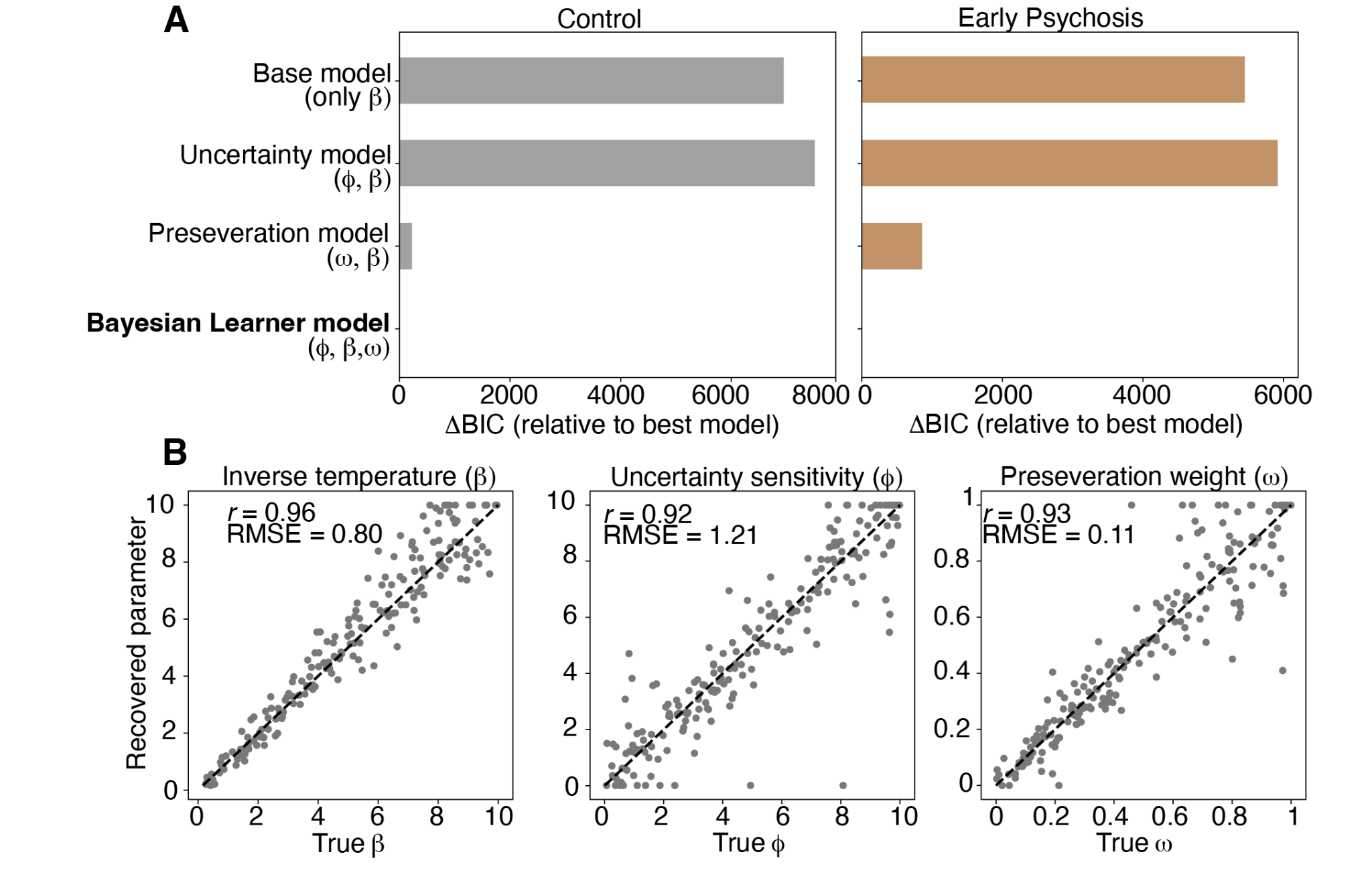

**Figure S4. Kalman Filter model ablation and comparison and parameter recovery for the Bayesian learner model. A**) Model comparison results shown as ΔBIC (relative to the best-fitting model) separately for Control and Early Psychosis (EP). Models compared include a base model containing only inverse temperature (β), an uncertainty model including uncertainty sensitivity and inverse temperature (φ, β), a perseveration model including perseveration weight and inverse temperature (ω, β), and the full Bayesian learner model including uncertainty sensitivity, inverse temperature, and perseveration (φ, β, ω). Lower ΔBIC values indicate better fit. The full Bayesian learner model provided the best fit in both groups. **B**) Parameter recovery analyses for the winning Bayesian learner model. Scatterplots show true versus recovered parameter values for inverse temperature (β), uncertainty sensitivity (φ), and perseveration weight (ω) across simulated datasets. Dashed lines indicate identity (y = x). Recovery was high for all parameters, indicating reliable parameter estimation under the model.

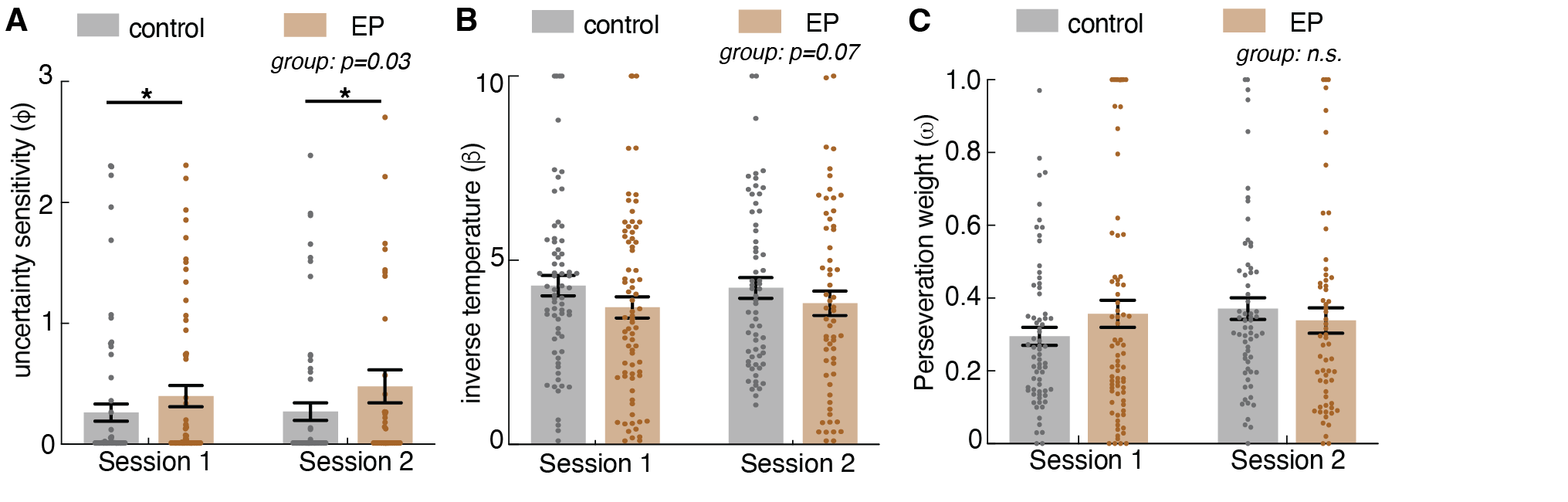

**Figure S5. Bayesian learner model parameters across Session 1 and Session 2. Related to Figure 3. A**) Uncertainty sensitivity (φ) fitted to participant choice sequences in Session 1 and Session 2. **B**) Inverse temperature (β) fitted to participants in Session 1 and Session 2. **C**) Perseveration weight (ω) fitted to participants in Session 1 and Session 2.

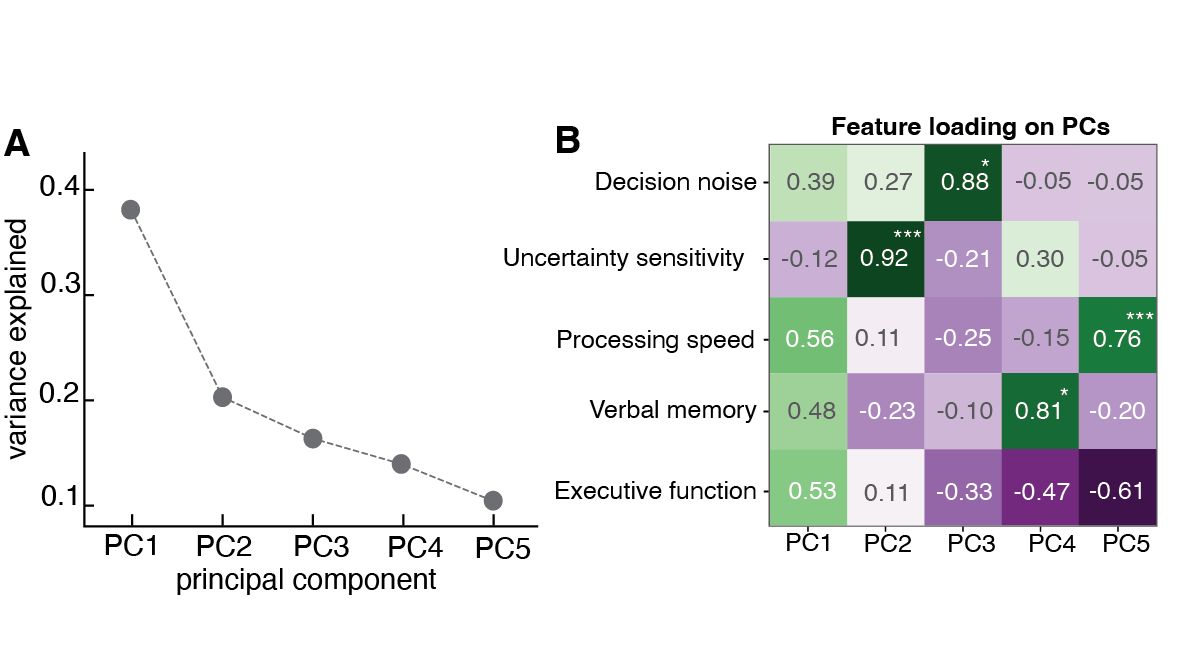

**Figure S6. Uncertainty sensitivity and decision noise are two independent microcognitive processes that influence explore-exploit tradeoff and are not captured with traditional measures of cognition among participants with early psychosis. A**) Variance in cognitive processing explained by five orthogonal principal components (PCs). The first three PCs capture the majority of the variance in cognitive processing (~75.4%). **B**) Feature loadings for each principal component. The computational parameters from the restless bandit task (Decision noise, Uncertainty sensitivity) captured unique variance from that of traditional cognitive measures (Processing Speed, Verbal Memory, Executive Function from Test My Brain cognitive battery). Significant feature loadings are indicated by * *p*<.05, ** *p*<.01, *** *p*<.001.

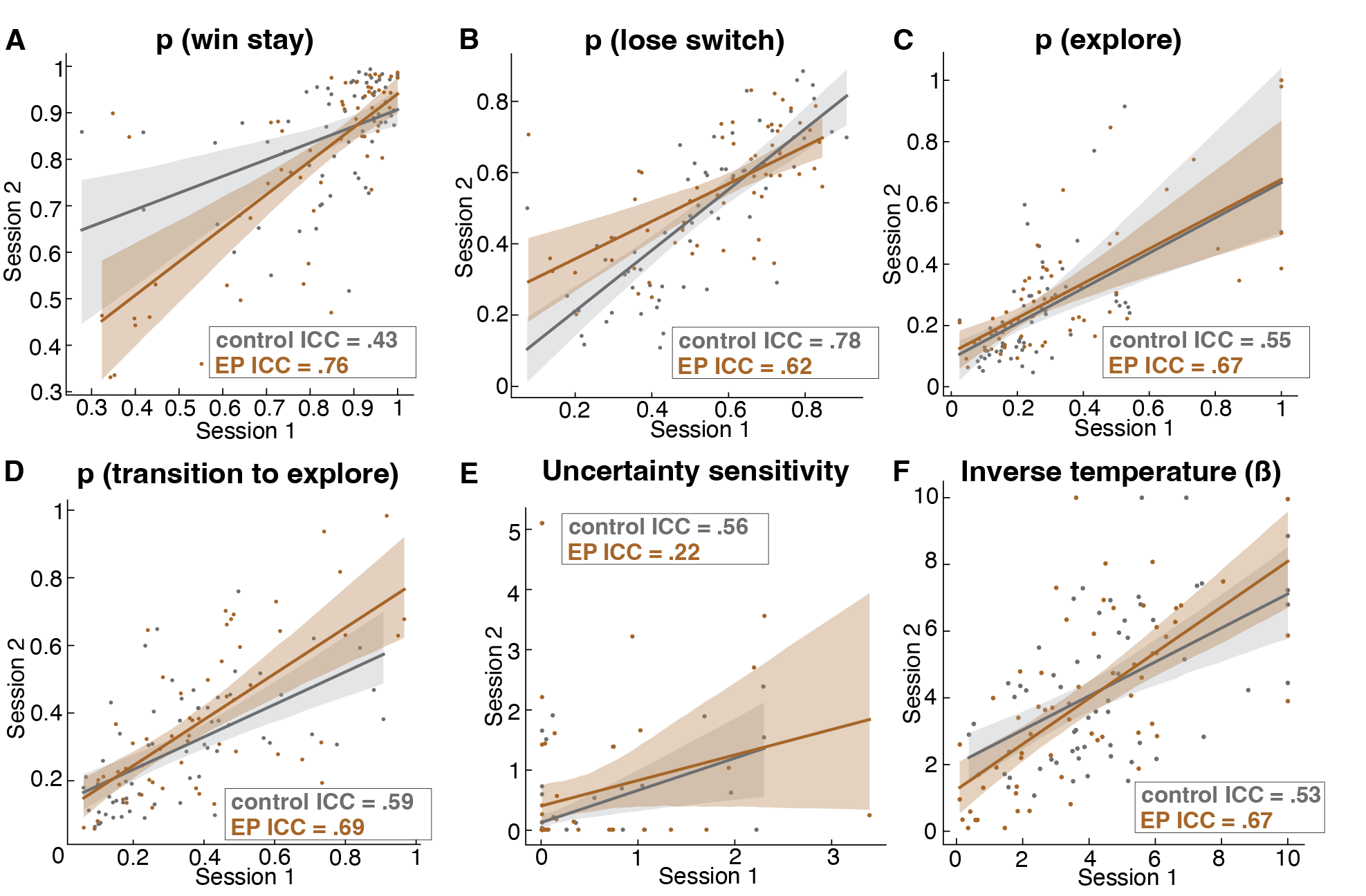

**Figure S7. Intraclass Correlation Coefficient (ICC) results revealed moderate test-retest reliability for model-free behavioral parameters and task computational parameters.** Task parameters showed moderate reliability using a two-way consistency ICC model for each participant group (control group, n=62, in black, Early Psychosis group, n=60, in blue) across session 1 and session 2, which occurred approximately 3 weeks apart. Each dot represents the correlation, across session 1 and session 2, between a single participant’s probability of engaging in the: **A**) explore strategy state, **B**) maintaining the exploit state strategy, **C**) win-stay strategy, **D**) lose-switch strategy, as well as each participant’s parameter values for: **E**) the Bayesian learner model-derived uncertainty sensitivity parameter, and **F**) the Bayesian learner model-derived decision noise parameter. The shaded area represents the 95% confidence interval around the regression line.

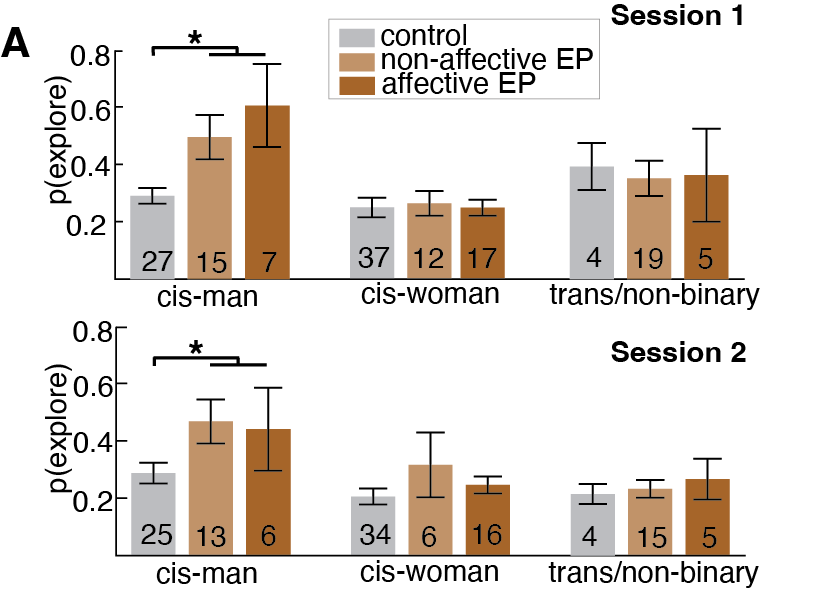

**Figure S8. Gender-by-diagnostic group analysis suggested that EP identifying as cis-men exhibited increased exploration.** Cis-men with non-affective psychosis and cis-men with affective psychosis had higher probability of exploration compared to controls in Session 1 and Session 2. * indicates p < 0.05, ** p<0.01, *** p <0.001. Graphs depict mean ± SEM across participants.

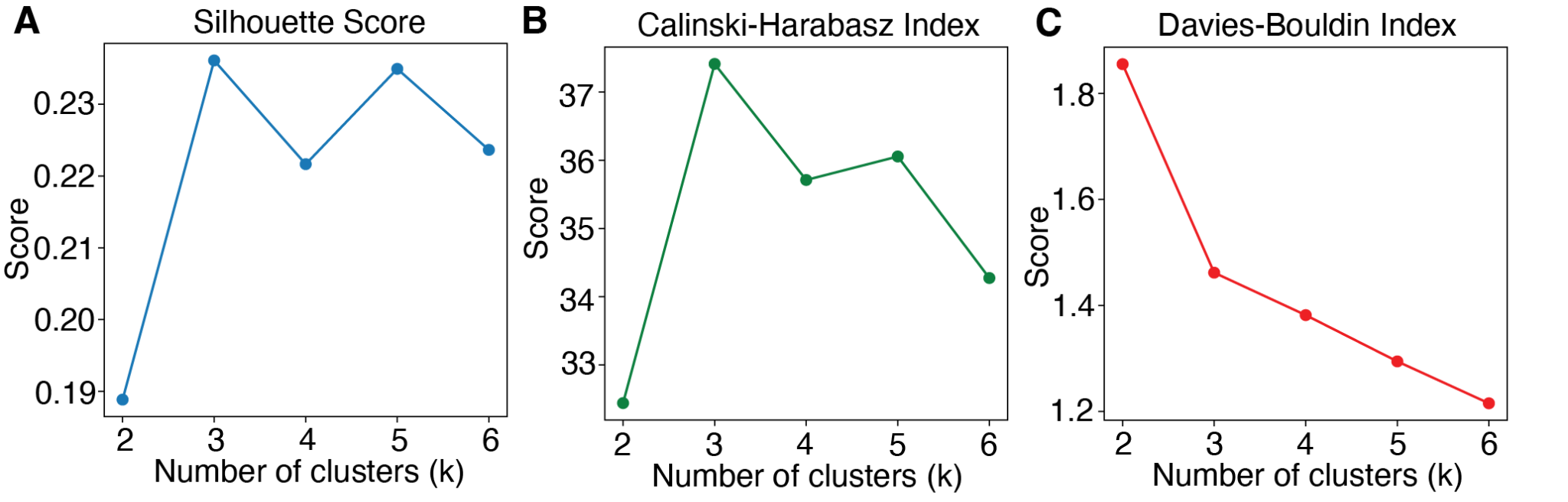

**Figure S9. Cluster metrics evaluating cluster separation, cohesion, and overall structure.**
